# Automated histopathological measurements of the tumor micro-environment predict distant metastasis after stage I/II Melanoma: discovery and validation in the population-based Dutch Early-Stage Melanoma (D-ESMEL) study

**DOI:** 10.64898/2026.06.02.26354705

**Authors:** Thamila Kerkour, Loes Hollestein, Alex Nigg, Yunlei Li, Jeffrey Damman, Catherine Zhou, Tamar Nijsten, Antien Mooyaart

**Author notes:** corresponding author: Antien Mooyaart, Department of Pathology & Clinical Bioinformatics, Dr. Molewaterplein 40, 3015GD Rotterdam. Funding sources: KWF Dutch Cancer Society. The funders of the study had no role in study design, data collection, data analysis, data interpretation, or the writing of the report. IRB approval status: The study was approved by the Erasmus MC Ethics Committee (MEC2020-0365). Patient consent: None to report.

## Abstract

**Background:** More than half of metastatic melanomas arise from patients initially diagnosed with early-stage melanoma. Objective biomarkers are needed to better identify high-risk patients.

**Objective:** To evaluate the prognostic value of multiple histopathological characteristics in predicting distant metastasis risk, in early-stage melanoma.

**Methods:** Using data from discovery set (n=442) and a population-based validation cohort (n=306, sampled from 5,815 patients) of the Dutch Early-Stage Melanoma (D-ESMEL) study, we investigated 14 histopathological characteristics of melanoma and their tumor micro-environment (TME) in an unprecedented integration, by expert pathologist scoring and automated quantitative measurements derived from a validated automated segmentation.

**Results:** Increased immune infiltrates (40% in cases vs. 50% in controls) were associated with lower risk of metastasis. Automated immune cell density was predictive in both the discovery set and the validation cohort, outperforming the manual pathological tumor infiltrating lymphocytes. The remaining histopathological features, including mitotic activity, did not retain independent value after controlling for current staging variables.

**Limitations:** TME evaluation in standard Hematoxylin-Eosin slides.

**Conclusion:** TME reaction is an important determinant of melanoma progression. The automated quantification of immune cell density appears to be a biomarker for distant metastasis risk. Further investigation into specific immune cell subtypes is required to facilitate clinical integration.

**Capsule summary:**

- In early-stage melanoma, automated quantification of immune cells in the tumor microenvironment predicts distant metastasis risk. Other histopathological features, including mitotic rate, lacked independent value after controlling for staging variables.
- These findings highlight the role of immune infiltrates in melanoma progression and support further research of specific immune cell subtypes.

## Introduction

Patients with early-stage cutaneous melanoma, classified as American Joint Committee on Cancer (AJCC) stage I or II, typically have a positive prognosis, but approximately 60% of all metastatic cases arise from these early-stage groups [1-3]. This clinical reality is explained by the high prevalence of stage I/II disease at diagnosis. Identifying biologically aggressive tumors within this large low-risk population is crucial. Novel biomarkers are needed to improve selection for (neo)adjuvant therapy and long-term outcomes. [4-7].

The current staging systems rely primarily on well-established histological features: Breslow thickness and ulceration [8, 9]. Additional tumor characteristics (e.g. mitotic rate and tumor-infiltrating lymphocytes) have been recognized for their potential prognostic value but it remains incompletely defined due to interobserver variability, and inconsistencies in their scoring [10-14]. Other histopathological features (e.g. hair involvement in the tumor and abnormal melanocyte migration within the epidermis) may further refine risk stratification but have not been systematically validated for their predictive potential [15-17].

Artificial intelligence (AI) is presented as a technology that will revolutionize clinical practice. The reality is that only about 1% of the proposed models ultimately achieve clinical implementation [18, 19]. This limitation is primary due to the lack of external validation and the challenges in integrating the tools into clinical pathological workflow [20-22]. As a result, AI remains a supportive tool rather than a routine clinical standard. We continue to rely on established histopathological features, where AI can play a valuable role by making certain histopathological features more quantifiable and therefore more objective.

In this study, we aim to determine whether a spectrum of histopathological features can identify early□stage melanoma patients at heightened risk of metastasis, independently of established staging variables. Using the largest population-based study worldwide of early-stage melanoma to study progression, including a discovery set and nationwide validation cohort of >5,800 patients: the Dutch Early-stage Melanoma (D-ESMEL) study [23, 24], we integrated automated quantitative analyses with expert histopathological assessment to enhance risk prediction for distant metastasis.

## Material and methods

### 1. Study design

The design of the D-ESMEL study has been previously described in detail [23, 24]. Briefly, the study includes a case-control discovery set matched on known prognostic features (i.e. Breslow thickness, ulceration, age and sex, n= 442) and a population-based validation cohort (n=5,815) with a nested case-control design, matched on AJCC stage (n=306) [24].

### 2. Histopathological scoring

Hematoxylin and eosin (H&E) slides of all excisions and biopsies were stained according to the manufacturer’s instructions using the HE600 (Ventana, Roche). All the slides were scanned and digitized at 40x magnification using the Hamamatsu Nanozoomer HT2.0 (Hamamatsu) [23, 24].

To determine which variables to score, we reviewed all the studies (2000-2021) on histological characteristics that may have a biological significance in melanoma progression (**Supplementary methods)**. Our final list included epidermal and architectural features (epidermal contour, pagetoid spread, nest formation and hair follicle involvement), tumor growth and characteristics (histological subtype, Clark level, mitotic index), tumor microenvironment (TME) and host response (tumor-infiltrating lymphocytes, plasma cell clusters and regression), patient context and secondary damage (pre-existing nevus, solar damage) (**Supplementary materials, Supplementary Table 1)**. Blinded to the outcome, a dermato-pathologist [AM] assessed digitalized H&E slides of the discovery set. In the validation cohort, the relevant features identified in the discovery set were re-assessed in 306 samples, in which 80 samples were independently scored by two pathologists [AM. JD]. Inter-observer agreement was assessed to assure that it was at least good (i.e. intraclass correlation or kappa >0.70) (**Supplementary methods)**.

### 3. Quantitative features extraction using automated model segmentation

We employed a validated deep learning model-based automated segmentation [25] to detect multiple histological features within the whole slide images (**Figure 1**). Segmentation quality control was performed as previously described [25]. We applied the model to both the discovery set and the validation cohort and generated quantitative measurements for each histological feature in only excision samples with adequate segmentation quality. Area, cell, and nuclei counting measurements were extracted. We extracted the area of the epidermis, the tumor microenvironment, hair follicles (within the whole tissue, in the TME), tumor nests and cells area. For cell-level measurements, we quantified the number of tumor cells, mitotic figures in the tumor area, and immune cells in the TME. To assess the relative immune response within the TME, the density of immune cells was calculated as a proportion relative to the total number of melanoma cells and immune cells together in the TME, rounded to the nearest decile (10% increments) as follows:

**Figure 1:**
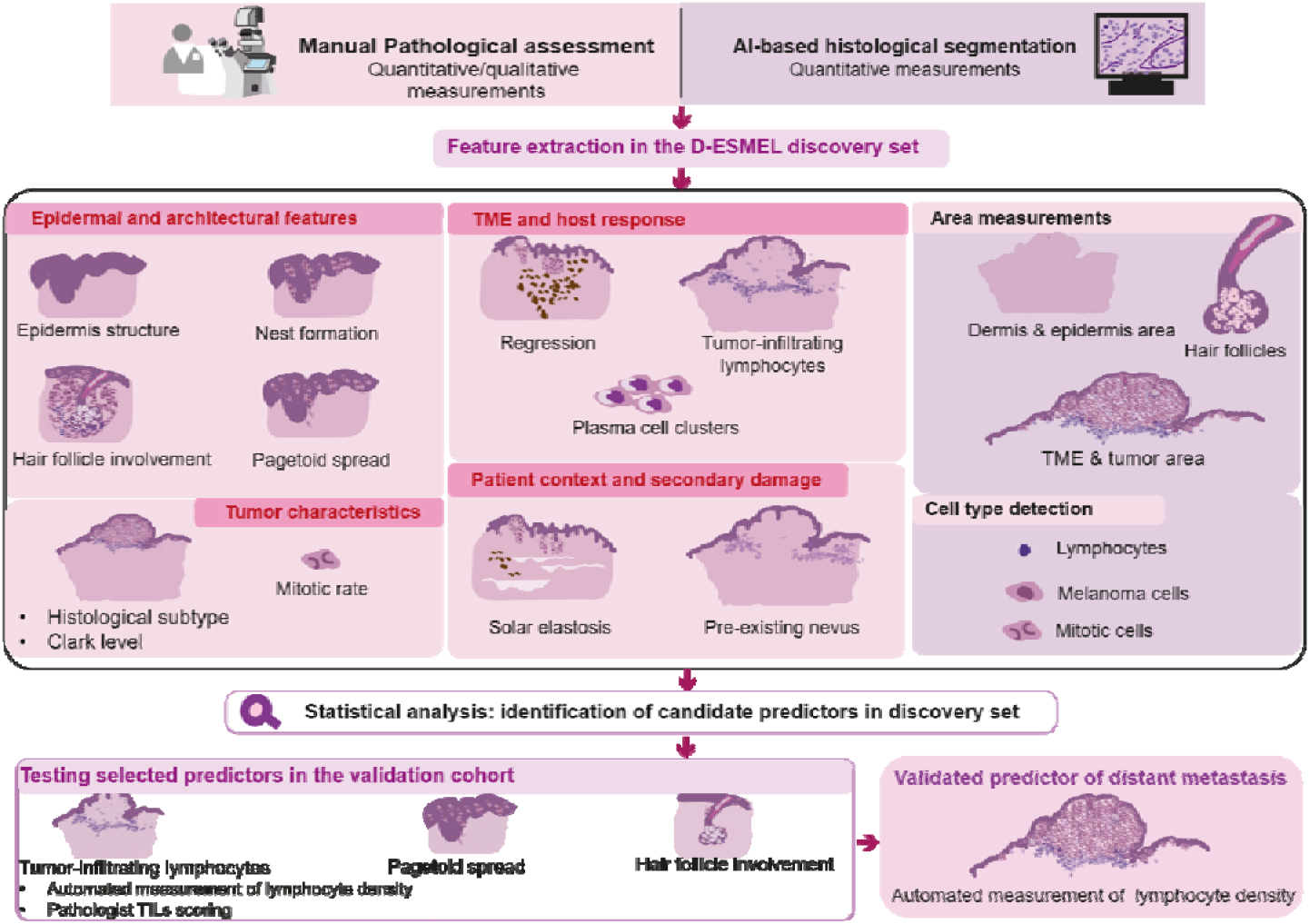
Overview of the experimental design and the key steps of the study.

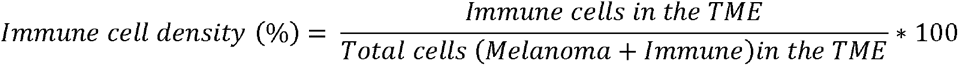

### 4. Manual versus automated pathological assessment

We evaluated the prognostic value of the pathologist-scored histological features and automated extracted features in two independent models to avoid collinearity. Agreement between approaches was assessed in the discovery cohort by correlating comparable features, using Spearman’s rank correlation (manual stromal TILs versus automated immune cell density, and mitotic rate versus automated mitotic counts).

### 5. Statistical analysis

The correlation between each histological feature and the risk of distant metastasis in the matched case-control sets were investigated using McNemar’s Chi-square for categorical variables, and Wilcoxon signed-rank test with continuity correction for continuous variables. In the discovery set, univariable and multivariable conditional logistic regression models were applied to assess associations with metastasis risk. Missing data in pathological scoring were assumed to be missing at random and handled using multiple imputations, using chained equation (*mice* R package (version 3.19.0) (**Supplementary Method)**. Backward selection was applied to identify independent predictors [26], (**Supplementary Method)**. Statistical interaction with AJCC stage was assessed by including an interaction term with statistically significant prognostic features and AJCC stage. Sensitivity analyses were performed to account for potential misclassification of sentinel lymph node biopsy status and to exclude matched sets in which a control patient developed a distant metastasis after their matched case’s metastasis date. Additionally, interaction analyses were performed to evaluate potential effect modification by stage, hair follicle involvement density combined with tumor location, and tumor subtype combined with the pagetoid spread, as it is a characteristic of superficial spreading melanomas [27].

Variables included in established risk prediction models from the Melanoma Institute Australia (MIA) were evaluated in multivariable models in the discovery set, as this was tightly matched on strong prognostic variables, thereby enabling to investigate additional prognostic value of other variables with maximum statistical power [28, 29] (**Supplementary method)**.

The prognostic value of identified features was evaluated in the validation cohort using conditional logistic regression, and their added value beyond the AJCC staging and the known clinicopathological features (age and sex) was assessed using Likelihood Ratio (LR) tests. Correlation between Breslow thickness (categorized as T1-T4) and mitotic activity (using both pathologist-reported mitotic rate and automated mitotic counts) was analyzed using Jonckheere-Terpstra trend and Pearson correlation tests. P values <0.05 were considered statistically significant. All analyses were performed using R version (4.4.0).

## Results

### 1. Patients’ characteristics

In the discovery set, 61% were male and the median age was between 62 and 64 years (**Supplementary Table 2**). Almost half of all samples were stage I melanoma (202/442, 46%).

In the validation cohort, 54% of all 153 cases were male and the median age was between 68 and 70 years. In this population-based sample, 56/153 (37%) of all distant metastasis after an early-stage melanoma occurred after an initial stage I melanoma. The distribution of patient and tumor characteristics was comparable between the complete datasets and the subsets of samples which were correctly automatically segmented (i.e. discovery dataset: 370/442 [84%]; validation cohort 238/306 [77%], **Supplementary Table 2**.).

### 2. Identification of the prognostic features in the discovery set

Of all manually scored variables, pagetoid spread occurred less frequently in the cases compared to the controls (OR = 0.58, 95% CI: 0.36-0.94), while hair follicles involvement in the tumor area was higher in cases compared to controls (OR= 1.56, 95% CI: 1.00-2.47) (**Table 1)**. All these variables remained significant in the multivariable model, while none of the other histopathological markers were different between cases and controls (**Table 2)**. In addition to our manually scored variables, prognostic variables from validated prediction models (e.g. histological subtype and location) did not have additional predictive value in addition to the matching variables in the discovery set (**Supplementary table 3)**, and therefore multivariable models were not further adjusted for those variables. Even mitotic rate did not show additional prognostic value, but mitotic activity increased with increased Breslow thickness, p-value <0.001) (**Supplementary figure 1A-D)**.

**Table 1:**
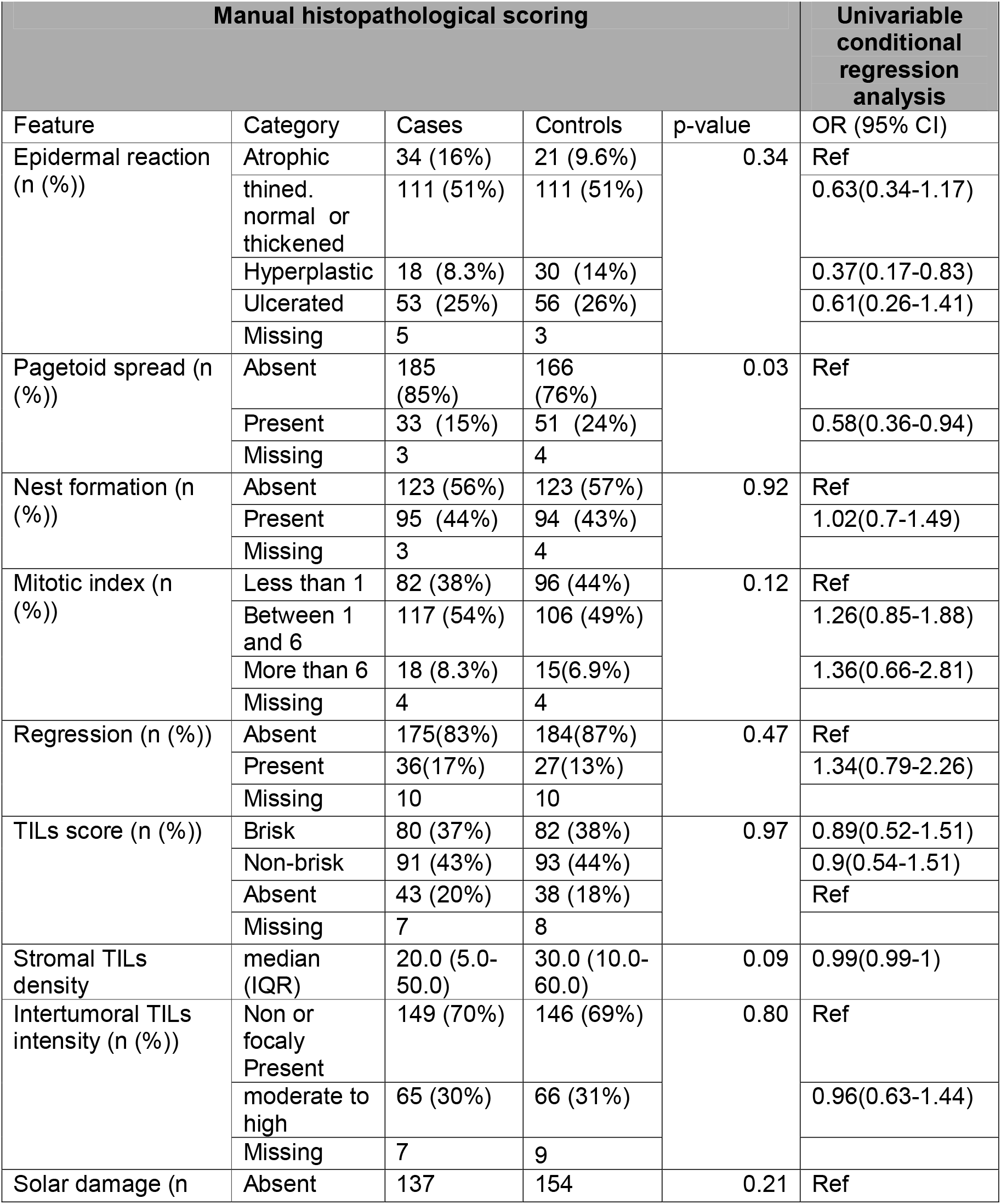
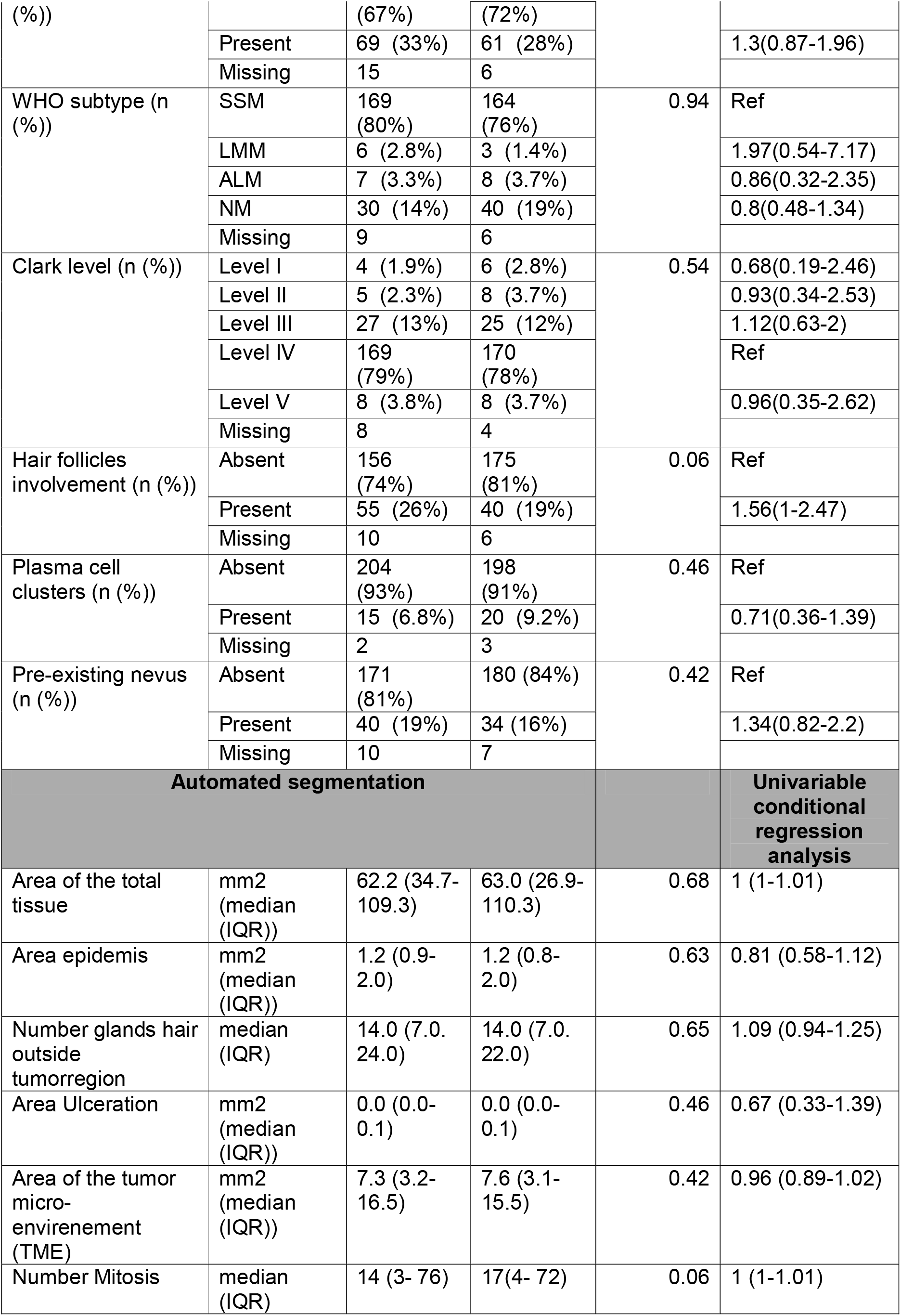
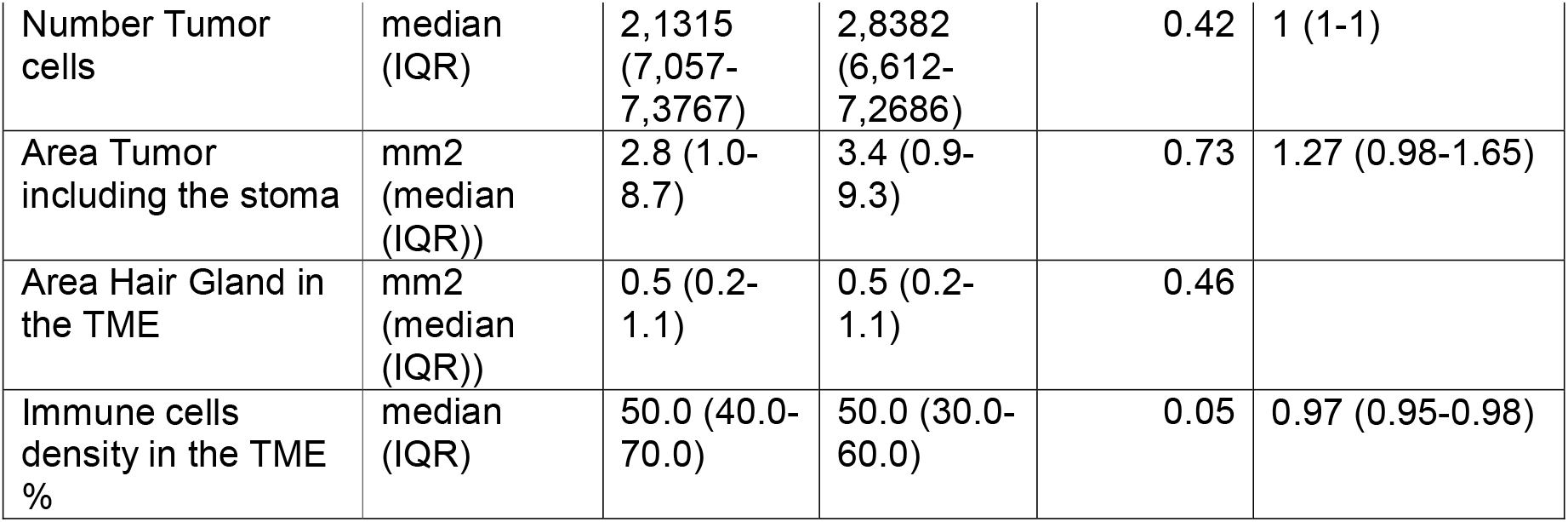
histopathological features, automated extracted measurements and conditional logistic regression to evaluate the association between the manual pathological measurements and the risk of distant metastasis on the discovery set of the D-ESMEL study. p-value was calculated using Wilcoxon signed rank test with continuity correction for continuous variables and McNemar’s Chi-squared test with categorical variables.

**Table 2:** Multivariable conditional logistic regression of the best predictors combination using the manual scoring or the automated detection.

| Manual histopathological scoring |  |  |
| --- | --- | --- |
| Variable | OR (95% CI) | p value |
| pagetoid spread if present | 0.36 (0.19-0.670) | 0.001 |
| Stromal TILs density (%) | 0.9 (0.84-0.98) | 0.01 |
| Hair follicles involvement if present | 2.04 (1.11-3.62) | 0.01 |
| Automated segmentation |  |  |
| Immune cells density in the TME in % | 0.97 (0.95-0.98) | 0.02 |

**Table 3:** Distribution of the best predictors in the validation cohort. p-value was calculated using Wilcoxon signed rank test with continuity correction for continuous variables and McNemar’s Chi-squared test for categorical variables.

|  |  | validation cohort (nested case-control study) |  | p-value |
| --- | --- | --- | --- | --- |
|  |  | Cases | Controls (matched on AJCC stage) |  |
| Manual histopathological scoring |  |  |  |  |
| pagetoid spread (n (%)) | Present | 34 (24%) | 35 (24%) | 0.78 |
|  | Missing | 11 | 7 |  |
| Stromal TILs density % | ( median (IQR) | 30 (10- 60) | 40 (20- 60) | 0.53 |
|  | Missing | 15 | 14 |  |
| Hair follicles involvement (n (%)) | Present | 42 (30%) | 35 (26%) | 0.33 |
|  | Missing | 16 | 11 |  |
| Automated segmentation |  |  |  |  |
| Immune cells density in the TME % | ( median (IQR) | 40(30-60) | 50 (30- 70) | 0.02 |

Pagetoid spread correlated with the superficial spreading subtype (Chi-square test, p-value <0.001), but there was no statistical interaction between pagetoid spread and the subtype (likelihood ratio test, p-value >0.05), (**Supplementary figure 2A)**. Similarly, hair follicle involvement correlated with the primary location of the tumor, (Chi-square test, p-value <0.0001), (**Supplementary figure 2C)**. However, no significant effect modification by tumor location on hair follicle involvement was observed when comparing models with and without the interaction term (likelihood ratio test, p = 0.97), therefore, only main effects of hair involvement were reported.

Of all automated segmentation measurements, immune cell density was predictive for distant metastasis (OR = 0.97 per 10% increase, 95% CI: 0.95-0.98) (**Table 2**). None other automated features were found to be predictive. A correlation (r=0.3, p-value<0.0001) was observed between manual stromal tumor-infiltrating lymphocytes (TILs) and automated immune cell density, as well as between mitotic rate and automated mitotic counts (r=0.7, p-value<0.0001) (**Supplementary figure 3A-B**). Similar patterns were observed, when accounting for sentinel lymph node biopsy, and excluding control patients who progressed after updating the follow-up (**Supplementary table 4)**.

### 3. Prognostic value of the histopathological variables in the validation cohort

The top-performing predictors identified in the discovery set were assessed in the validation cohort. The manual histopathological variables were not validated, but the automated segmentation measure of immune cell density in the TME was prognostic (**Table 4**). A higher immune cell density was associated with lower risk of distant metastasis (adjusted OR per percentage 10% increase in immune cells in the TME: adjusted OR = 0.98 [95%CI: 0.97-0.99].

**Table 4:**
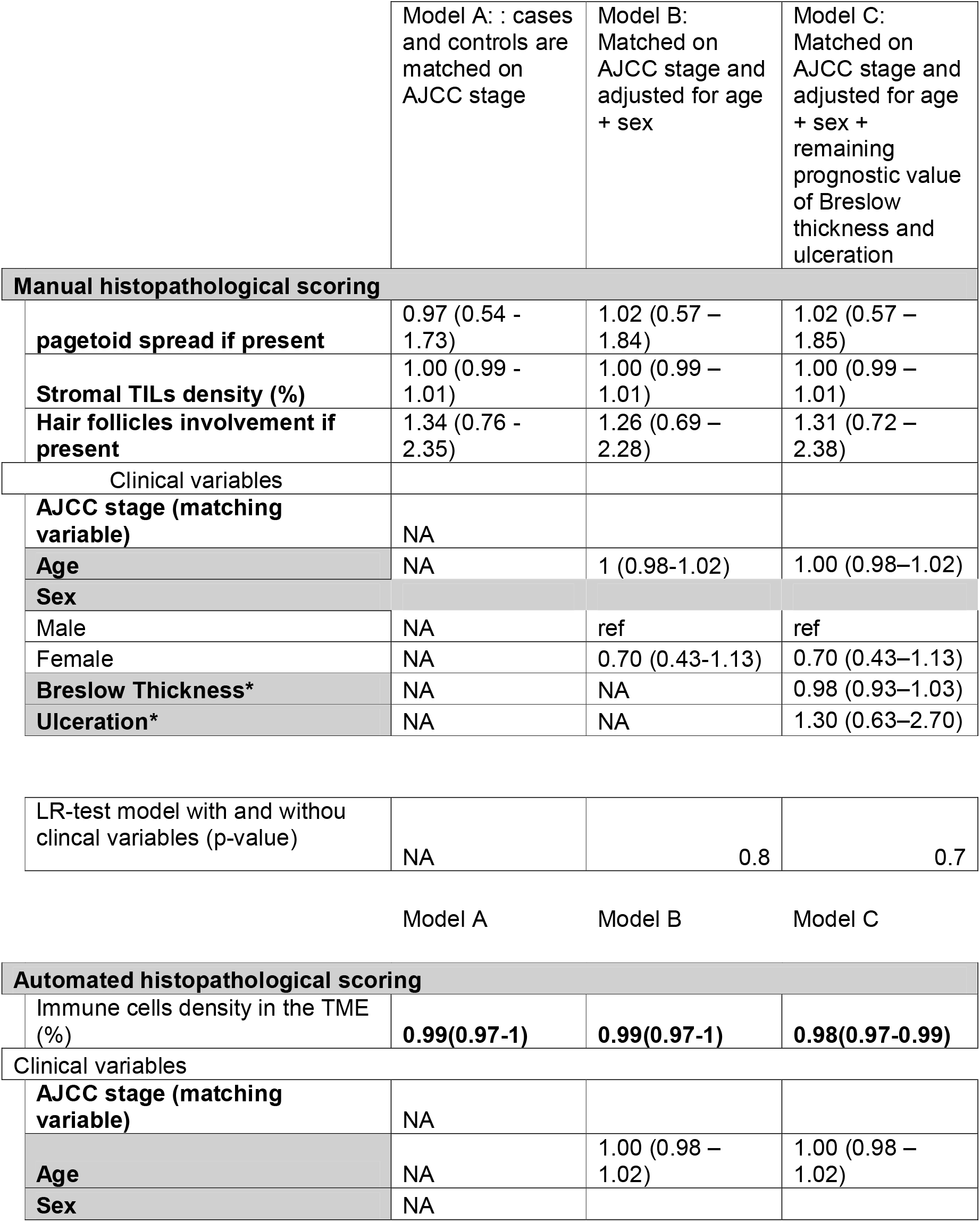
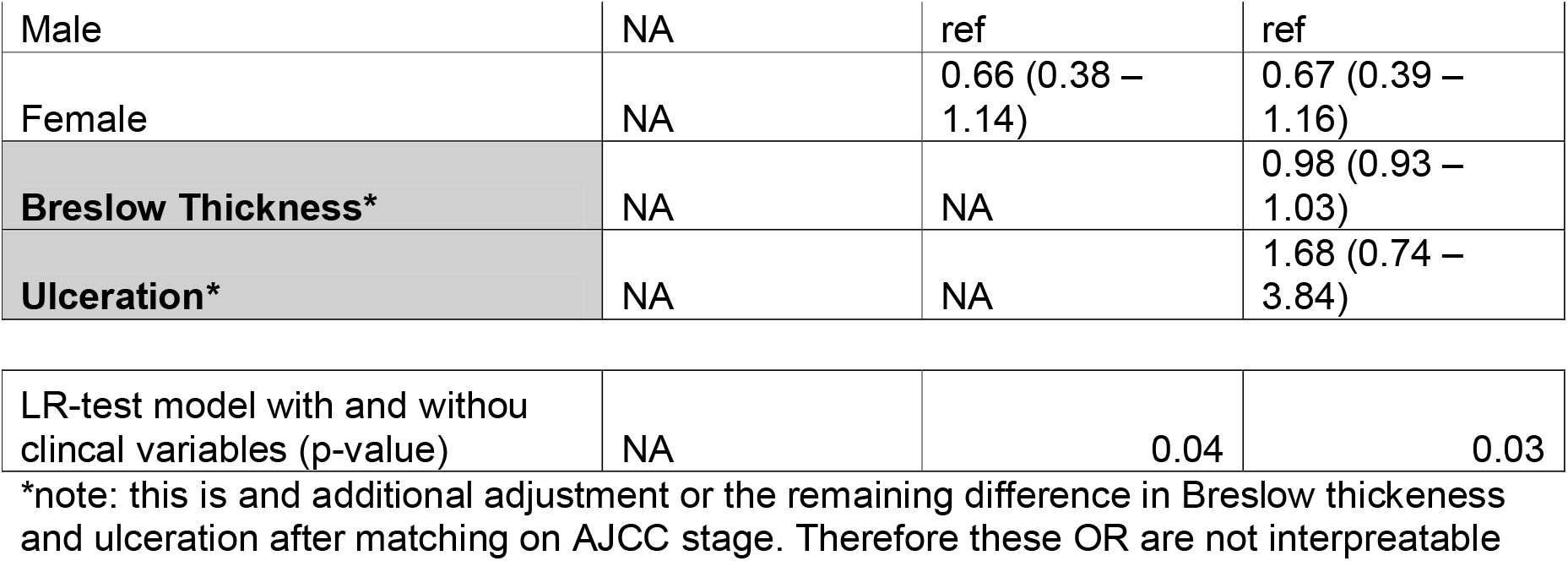
Validation of the prognostic histological features in the nested case-control validation cohort and testing for the improvement of the currents models.

|  | Model A: : cases and controls are matched on AJCC stage | Model B: Matched on AJCC stage and adjusted for age + sex | Model C: Matched on AJCC stage and adjusted for age + sex + remaining prognostic value of Breslow thickness and ulceration |
| --- | --- | --- | --- |
| **Manual histopathological scoring** |  |  |  |
| **pagetoid spread if present** | 0.97 (0.54 - 1.73) | 1.02 (0.57 – 1.84) | 1.02 (0.57 – 1.85) |
| **Stromal TILs density (%)** | 1.00 (0.99 - 1.01) | 1.00 (0.99 – 1.01) | 1.00 (0.99 – 1.01) |
| **Hair follicles involvement if present** | 1.34 (0.76 - 2.35) | 1.26 (0.69 – 2.28) | 1.31 (0.72 – 2.38) |
| Clinical variables |  |  |  |
| **AJCC stage (matching variable)** | NA |  |  |
| **Age** | NA | 1 (0.98-1.02) | 1.00 (0.98–1.02) |
| **Sex** |  |  |  |
| Male | NA | ref | ref |
| Female | NA | 0.70 (0.43-1.13) | 0.70 (0.43–1.13) |
| **Breslow Thickness\*** | NA | NA | 0.98 (0.93–1.03) |
| **Ulceration\*** | NA | NA | 1.30 (0.63–2.70) |
| LR-test model with and without clinical variables (p-value) | NA | 0.8 | 0.7 |
| **Automated histopathological scoring** |  |  |  |
| Immune cells density in the TME (%) | **0.99(0.97-1)** | **0.99(0.97-1)** | **0.98(0.97-0.99)** |
| Clinical variables |  |  |  |
| **AJCC stage (matching variable)** | NA |  |  |
| **Age** | NA | 1.00 (0.98 – 1.02) | 1.00 (0.98 – 1.02) |
| **Sex** | NA |  |  |
| Male | NA | ref | ref |
| Female | NA | 0.66 (0.38 – 1.14) | 0.67 (0.39 – 1.16) |
| **Breslow Thickness\*** | NA | NA | 0.98 (0.93 – 1.03) |
| **Ulceration\*** | NA | NA | 1.68 (0.74 – 3.84) |
| LR-test model with and without clinical variables (p-value) | NA | 0.04 | 0.03 |

\*note: this is and additional adjustment or the remaining difference in Breslow thickness and ulceration after matching on AJCC stage. Therefore these OR are not interpretable

Comparable to the discovery set, in the validation cohort we observed correlation between pagetoid spread and histological subtype, and between hair follicles involvement and location, Chi-square test, p-value <0.001, (**Supplementary figure 2B**, and **2D** respectively). Interaction between location and hair follicle involvement was statistically significant (p-value = 0.05), indicating differential effects on different locations (**Supplementary table 5)**.

## Discussion

Using the largest stage I/II melanoma study to study progression (D-ESMEL), we observed that immune-infiltrating lymphocytes are protective against progression to distant metastasis. Patients without distant metastasis presented higher immune infiltrates compared to cases, suggesting that the host immune response is an independent determinant of melanoma progression.

By systematically assessed immune cells using both conventional and automated approaches, we demonstrated that quantitative immune density in the TME, rather than the qualitative assessment, provided a reproducible prognostic signal across both the discovery set and the validation cohort. Several studies reported the association of TILs scoring with improved survival in melanoma [10, 14, 30, 31]. For instance, recent work by Adigbli and colleagues demonstrated that Brisk scoring improved the risk stratification in stage II melanoma [31]. Although, previous research highlight that independent prognostic value of TILs remains debated due to their heterogeneous scoring and interobserver variability [32, 33]. Our findings align with these observations and show that the prognostic signal of the immune cells is more reliably captured through an objective measurement. Importantly, our results support that the TME provides a prognostic value beyond established staging variables. However, the true predictive improvement remains moderate and does not yet justify it implementation in current clinical risk models. More refined biomarkers, such as detailed characterization of immune cell subtypes, are likely required before characteristics of the TME can be effectively integrated into clinical staging system.

Most of the manually scored histological characteristics were not prognostic after strong matching on Breslow thickness, ulceration, sex and age. Several features have previously been reported as predictors of metastasis progression, including mitotic rate, regression, solar damage, and Clark level [28, 29, 34, 35]. This contradictory observation can be explained by the fact that the previous research was based on clinical cohorts that overrepresented aggressive cases or did not sufficiently correct for staging variables. Here, we show that when the study design is strictly controlled for Breslow thickness, ulceration, age, and sex, most secondary histological variables have no independent prognostic value. Our analysis suggests that many reported associations likely reflect their correlation with tumor thickness, rather than independent biological effects on tumor malignancy and progression. Particularly, mitotic rate does not provide additive value on top of Breslow thickness, although it was previously incorporated into staging criteria [36]. From a biological perspective, increased mitotic activity contributes to tumor growth and therefore increased tumor thickness. As a result, the proliferative behavior of the tumor may already be captured by the vertical growth reflected in Breslow thickness. This relationship explains why the independent prognostic contribution of mitotic rate becomes insignificant once tumor thickness is accounted for.

Besides tumor immune reaction, we obtained discrepant findings regarding the pagetoid spread and hair follicle involvement. Pagetoid spread decreased the metastasis risk in the discovery set but was not replicated in the validation cohort. This finding may be due to chance rather than representing a true prognostic variable or due to the interobserver variability between our two pathologists. Alternative explanation consists of the matching strategy, in which superficial spreading melanomas (case) with less pagetoid spread may have been matched with a control with a potentially more aggressive subtype of similar thickness. Similarly, hair follicle involvement did not retain prognostic value in the validation cohort. Although, exploratory analyses suggested a location-dependent effect. Therefore, follicular involvement may not warrant inclusion in general population risk calculators, but possibly, it could be considered a high-risk morphological feature in the specific context of non-chronic sun-damaged skin.

Our study has several notable strengths. First, the unique design of the D-ESMEL study facilitates the evaluation of histological features with sufficient metastatic events in stage I/II melanoma and subsequently facilitates validation of the prognostic value in a population-based cohort. Secondly, we provided a detailed description of the scored features and used a validated automated segmentation method [25], thereby increasing the reproducibility. Thirdly, we combined both the expert pathological measurements and automated quantification analysis, which allowed us to compare human scoring to objective measurements. Together, our work provides a uniquely comprehensive answer to the longstanding debates surrounding histological biomarkers of melanoma progression.

Limitations of our study include the low number or rare melanoma subtypes and the quantification of the immune cell density based on H&E slide, which does not capture the special immune organization and functionalities such as the T cell activation states. To address this issue, multiplex immunofluorescent studies investigate the role of tumor-immune interaction are needed.

In conclusion, our study showed that the TME is an independent determinant of distant metastasis in early-stage melanoma. Automated immune cell density measurements outperformed manual immune cell assessments. These results highlight the potential value of integrating quantitative digital pathology approaches into melanoma risk stratification and support further investigation of immune-based biomarkers for improving prognostic assessment in early-stage disease.

## Supporting information

all supplementary tables and figures

## Data Availability

All data produced in the present study are available upon reasonable request to the authors

## Supplementary tables and figures titles

- Supplementary table 1: Descriptive table of the manual histopathological measurements scored by pathologists.
- Supplementary Table 2 : Clinical data for patients used in Manual histopathological scoring and automated segmentation method : Discovery set and the nested case-control validation cohort with randomly selected controls.
- Supplementary table 3: MIA histopathological variables prediction in the D-ESMEL study, * p value was calculated using the Likelihood ratio test for categorical variables with more than 2 categories. n.i: not included in the model.
- Supplementary Table 4: Subgroup analysis in the discovery set for the best combination predictors.
- Supplementary table 5: Interaction term between hair follicle and location in the validation cohort.
- Supplementary figure 1: Mitotic index concordance with the tumor thickness in discovery set and validation cohort.
- Supplementary figure 2: Hair involvement across the location, and pagetoid spread across histological subtype in discovery set and validation cohort.
- Supplementary figure 3: Comparison between AI and pathologist based measurements

## Notes

Conflicts of Interest: All the authors of this study, have no conflicts of interest to disclose.

### Competing Interest Statement

The authors have declared no competing interest.

### Author Declarations

The study was approved by the Erasmus MC Ethics Committee (MEC2020-0365)

