## Supplementary material for "Automated histopathological measurements of the tumor micro-environment predict distant metastasis after stage I/II Melanoma: discovery and validation in the population-based Dutch Early-Stage Melanoma (D-ESMEL) study": all supplementary tables and figures: Supplementary materials.pptx

### Slide 1
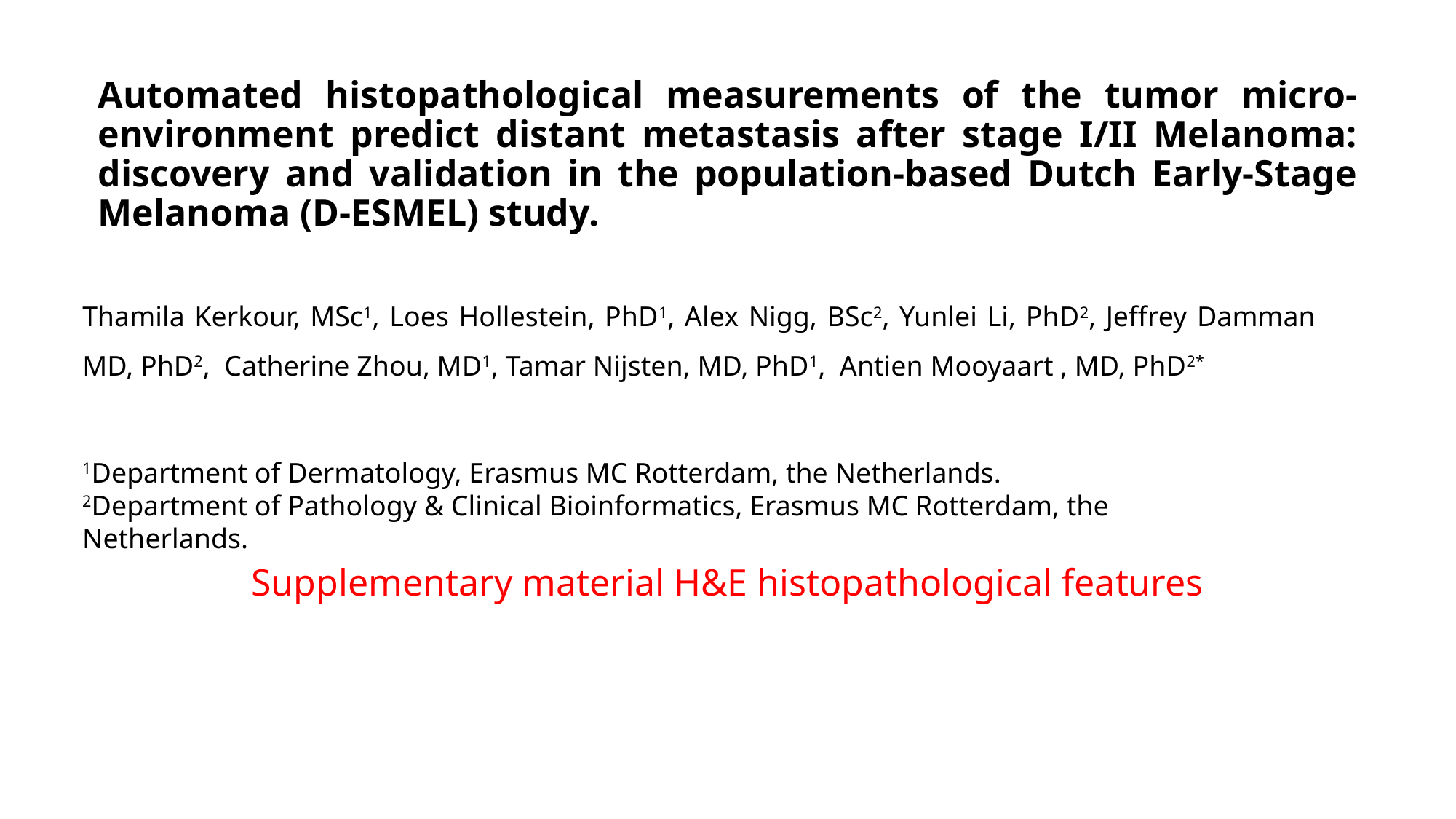

Automated histopathological measurements of the tumor micro-environment predict distant metastasis after stage I/II Melanoma: discovery and validation in the population-based Dutch Early-Stage Melanoma (D-ESMEL) study.
Thamila Kerkour, MSc^1^, Loes Hollestein, PhD^1^, Alex Nigg, BSc^2^, Yunlei Li, PhD^2^, Jeffrey Damman MD, PhD^2^, Catherine Zhou, MD^1^, Tamar Nijsten, MD, PhD^1^, Antien Mooyaart , MD, PhD^2*^
^1^Department of Dermatology, Erasmus MC Rotterdam, the Netherlands.
^2^Department of Pathology & Clinical Bioinformatics, Erasmus MC Rotterdam, the Netherlands.
#### Supplementary material H&E histopathological features

### Slide 2
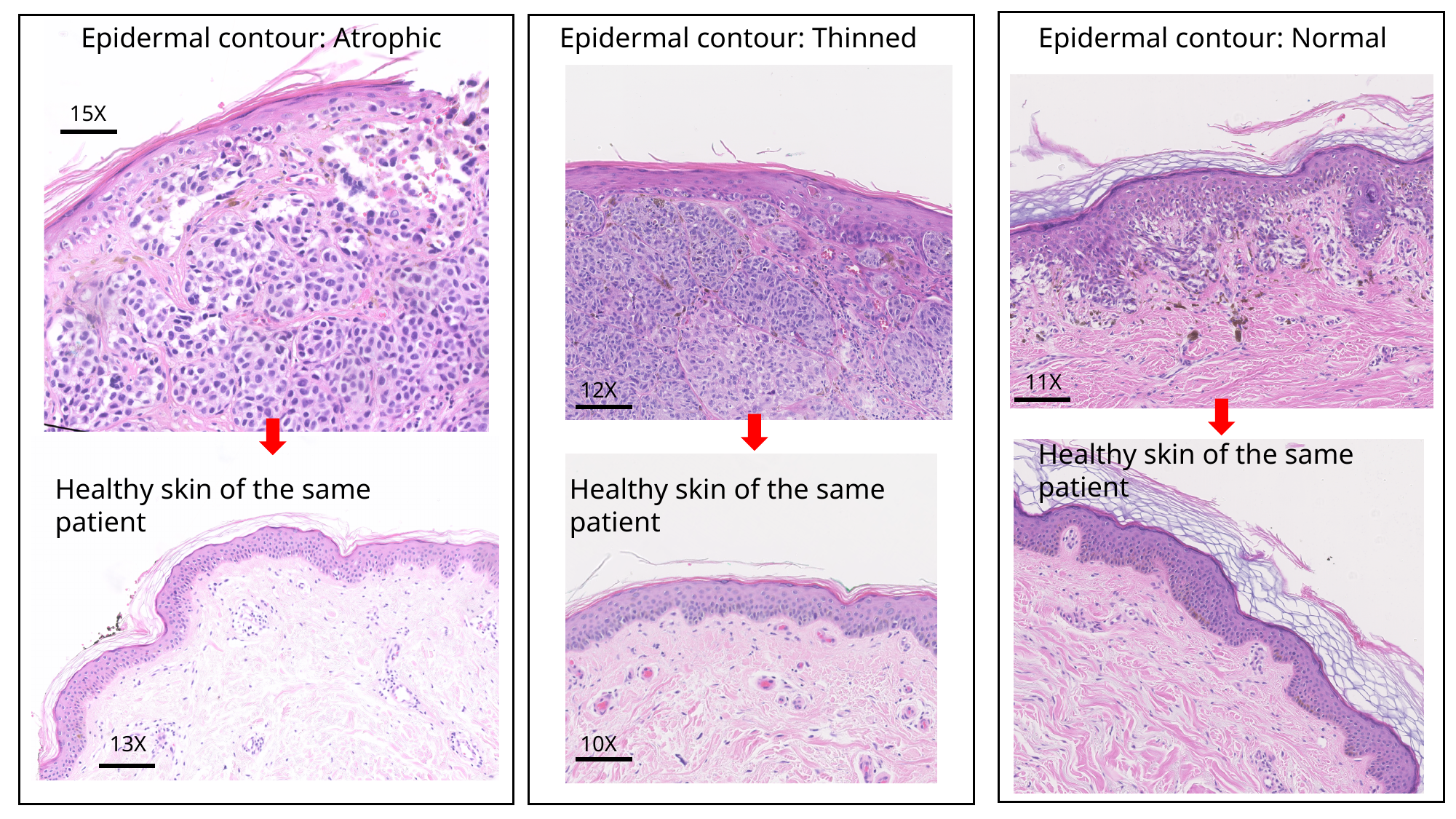

Epidermal contour: Atrophic
15X
Epidermal contour: Thinned
Epidermal contour: Normal
13X
11X
12X
Healthy skin of the same patient
Healthy skin of the same patient
13X
Healthy skin of the same patient
10X

### Slide 3
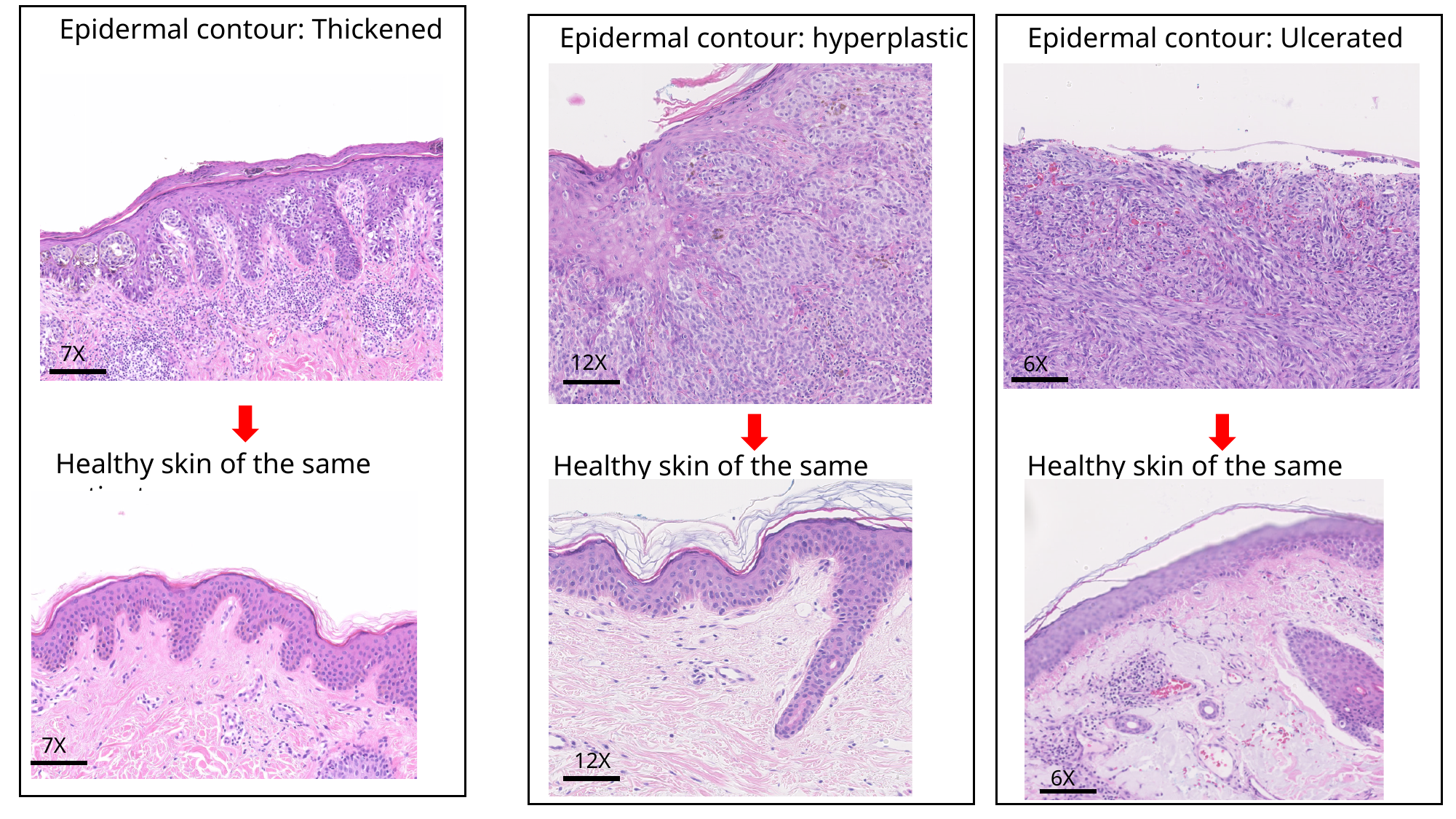

Epidermal contour: Thickened
Epidermal contour: hyperplastic
Epidermal contour: Ulcerated
7X
12X
6X
Healthy skin of the same patient
Healthy skin of the same patient
Healthy skin of the same patient
7X
12X
6X

### Slide 4
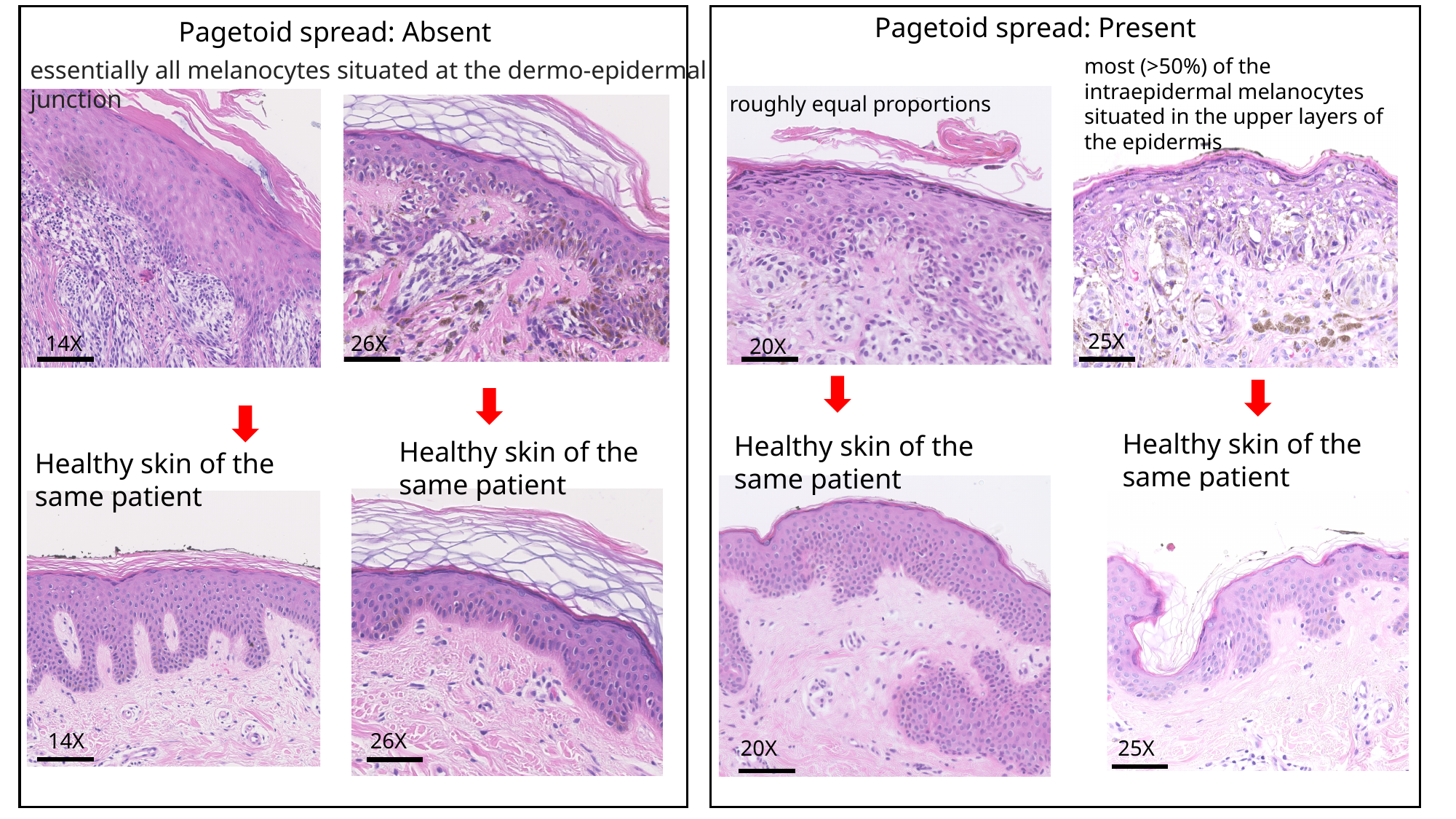

Pagetoid spread: Present
Pagetoid spread: Absent
most (>50%) of the intraepidermal melanocytes situated in the upper layers of the epidermis
essentially all melanocytes situated at the dermo-epidermal junction
roughly equal proportions
25X
14X
26X
20X
Healthy skin of the same patient
Healthy skin of the same patient
Healthy skin of the same patient
Healthy skin of the same patient
26X
14X
25X
20X

### Slide 5
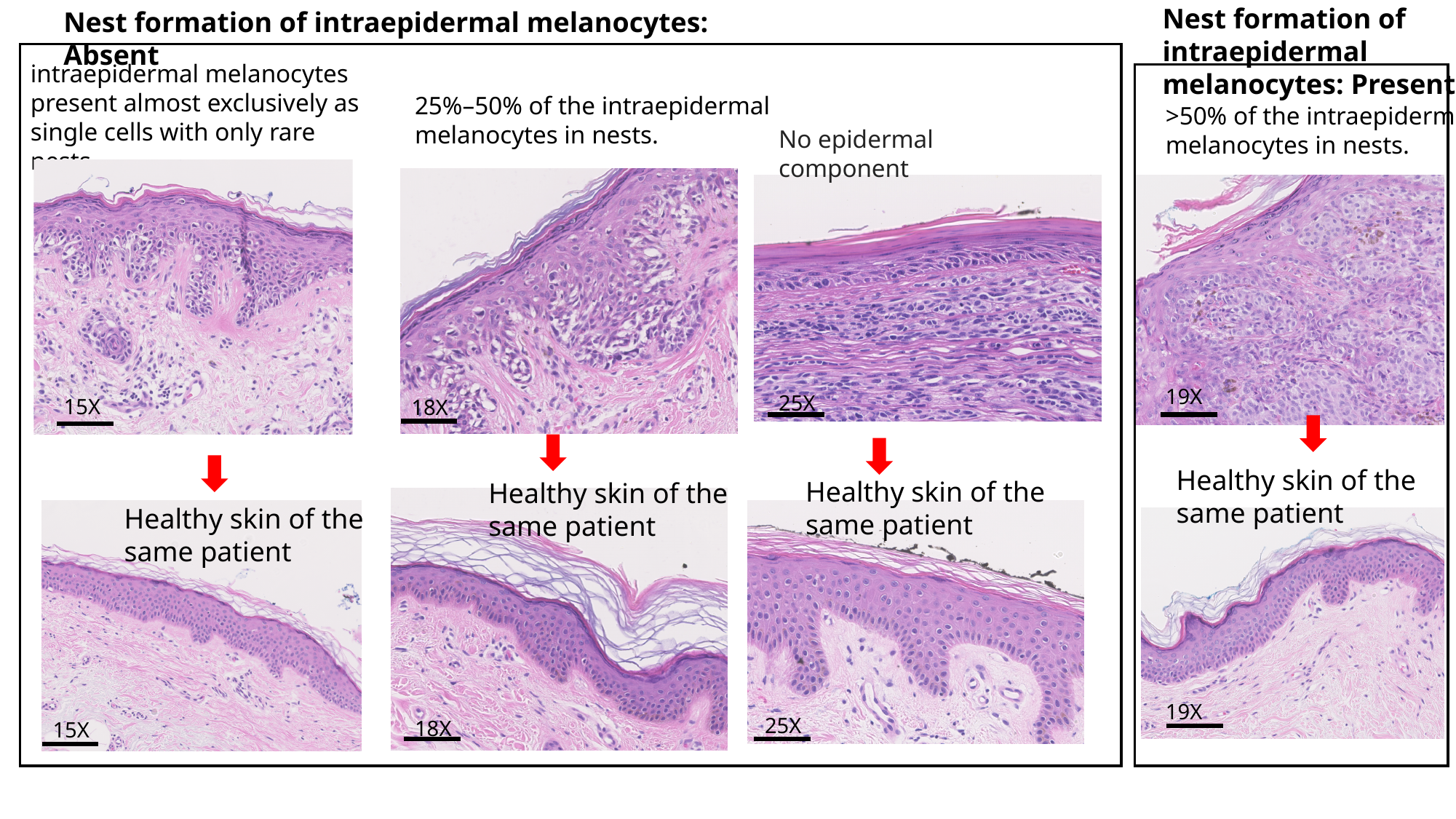

Nest formation of intraepidermal melanocytes: Absent
Nest formation of intraepidermal melanocytes: Present
intraepidermal melanocytes present almost exclusively as single cells with only rare nests.
25%–50% of the intraepidermal melanocytes in nests.
>50% of the intraepidermal melanocytes in nests.
No epidermal component
19X
25X
15X
18X
Healthy skin of the same patient
Healthy skin of the same patient
Healthy skin of the same patient
Healthy skin of the same patient
19X
25X
18X
15X

### Slide 6
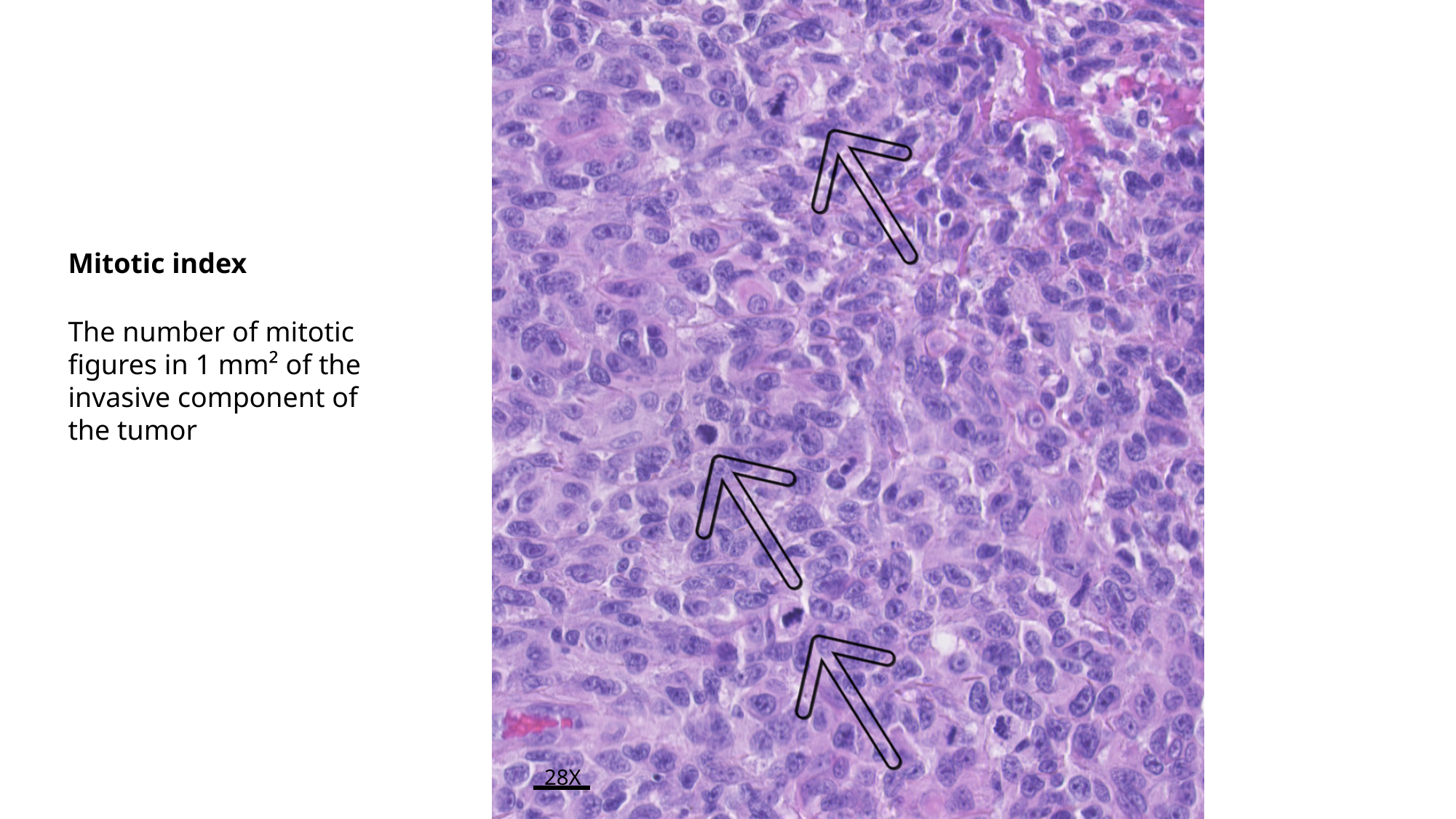

Mitotic index
The number of mitotic figures in 1 mm² of the invasive component of the tumor
28X

### Slide 7
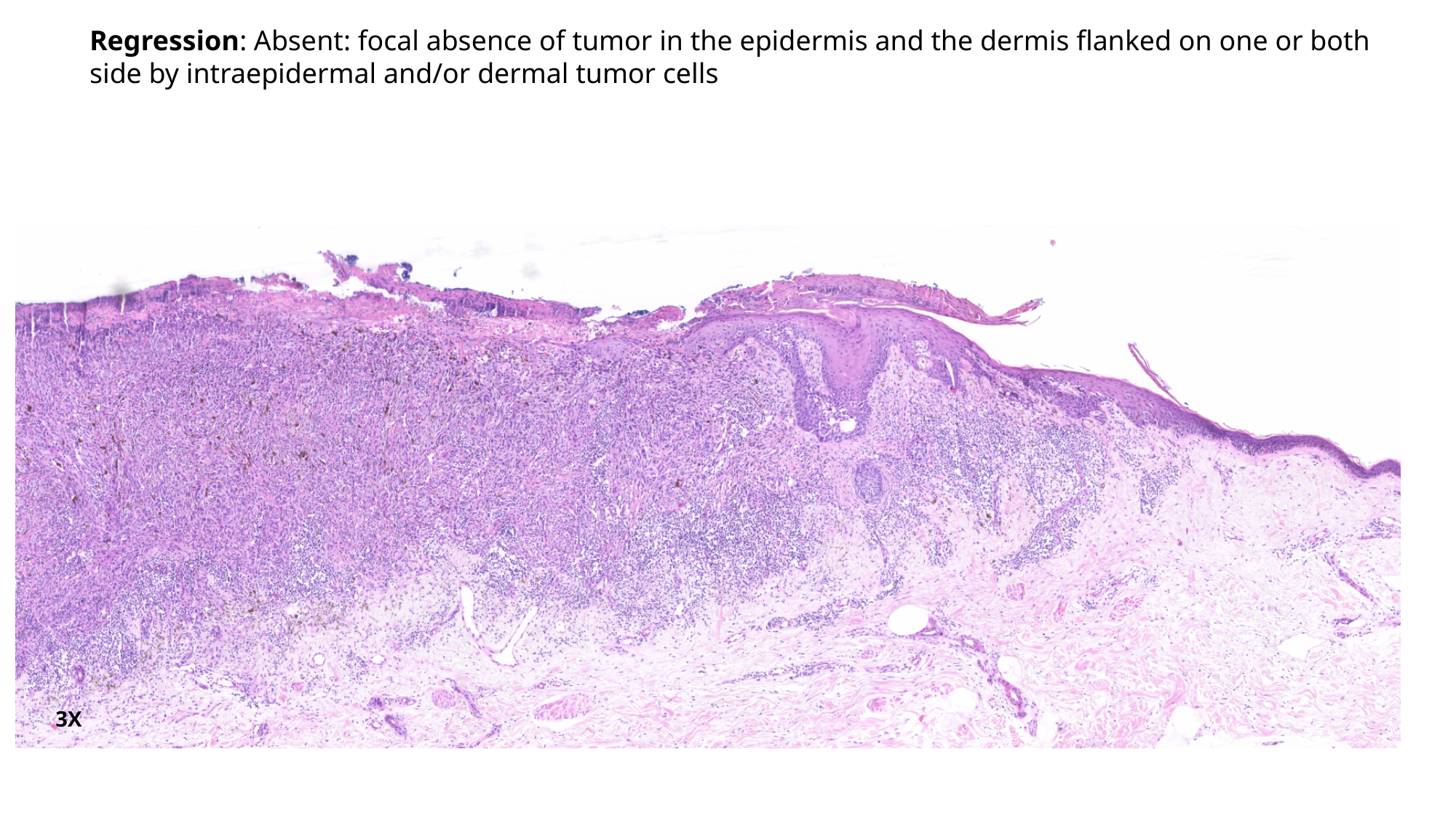

Regression: Absent: focal absence of tumor in the epidermis and the dermis flanked on one or both side by intraepidermal and/or dermal tumor cells
3X

### Slide 8
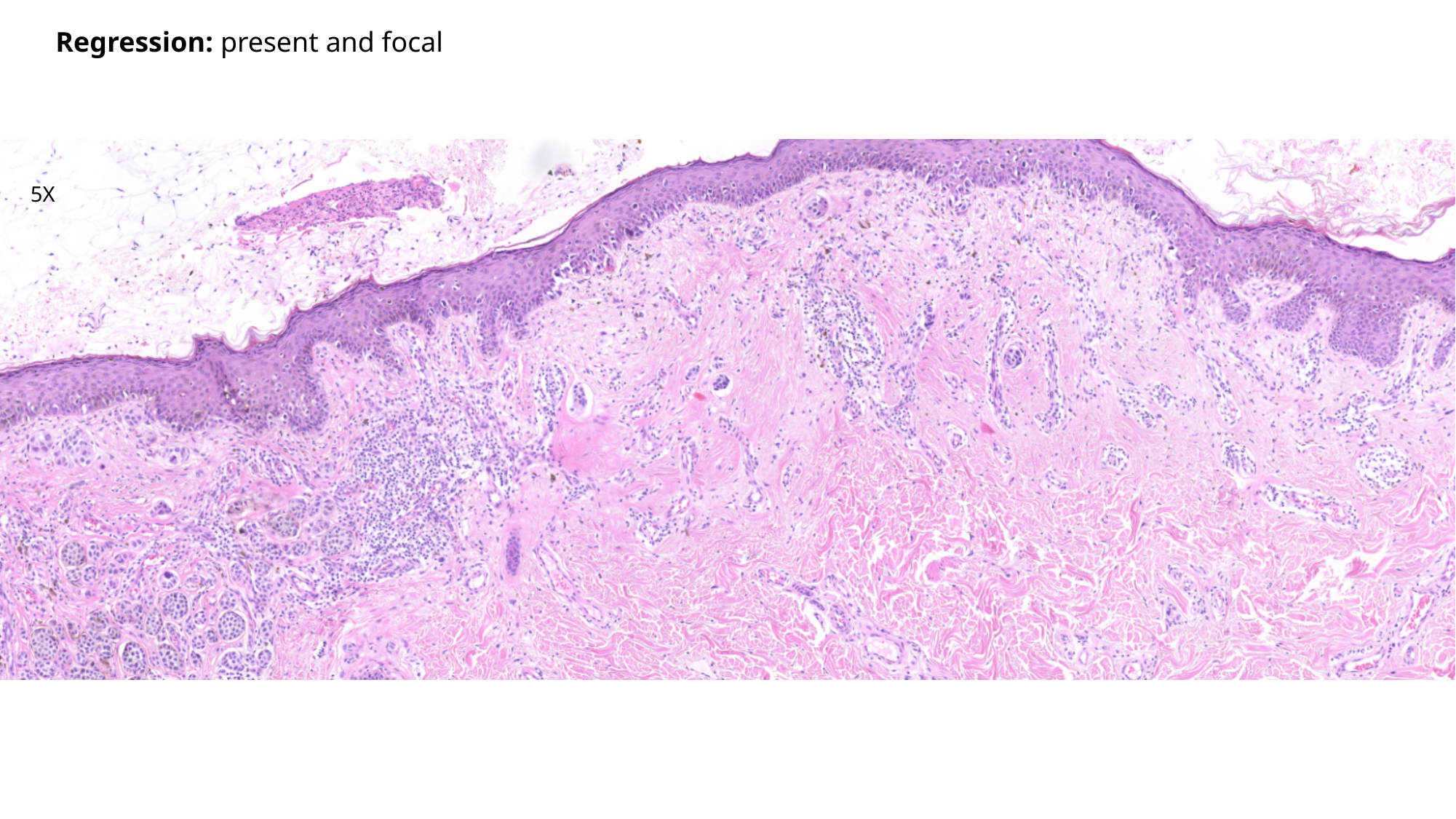

Regression: present and focal
5X

### Slide 9
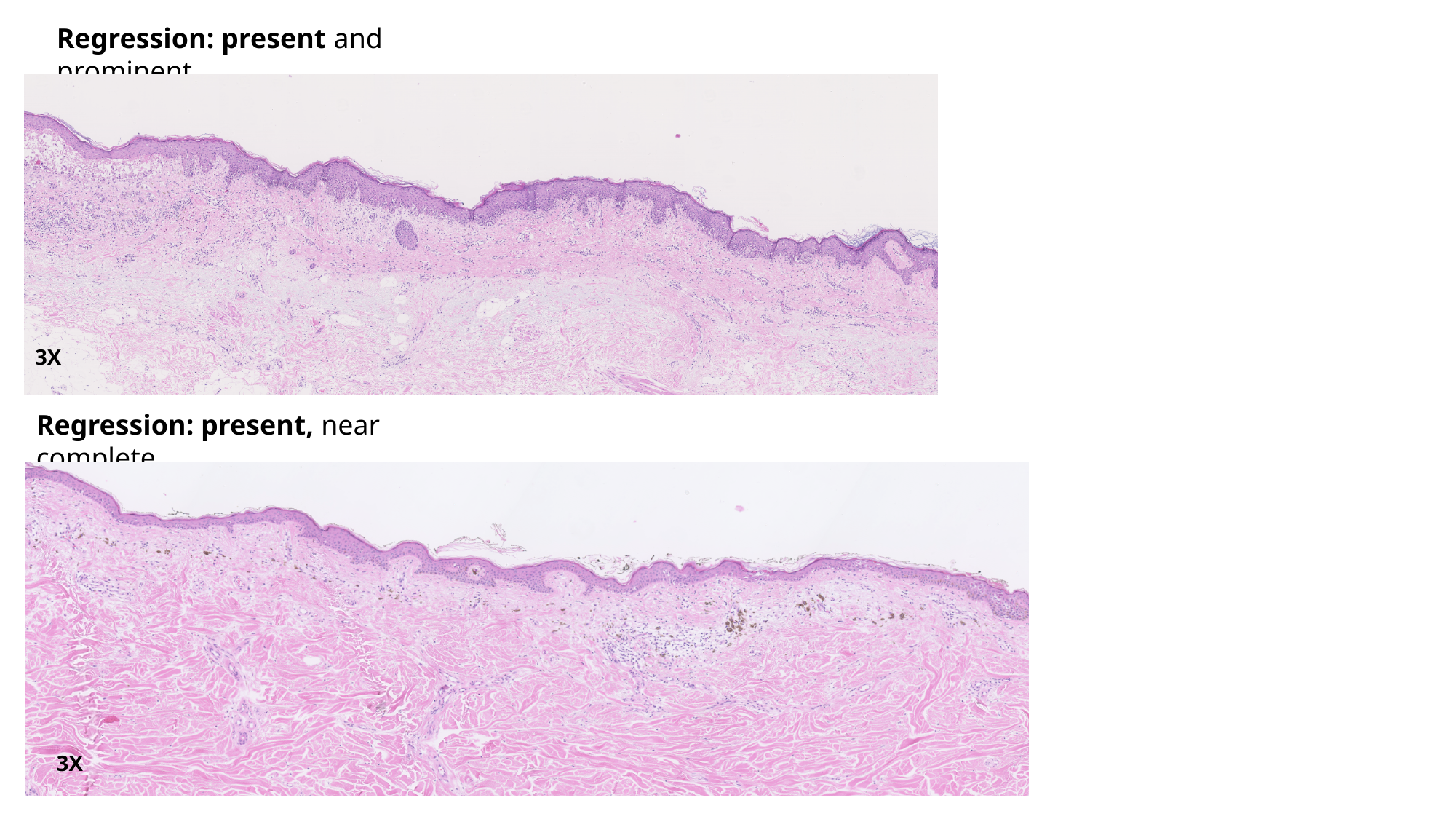

Regression: present and prominent
3X
Regression: present, near complete
3X

### Slide 10
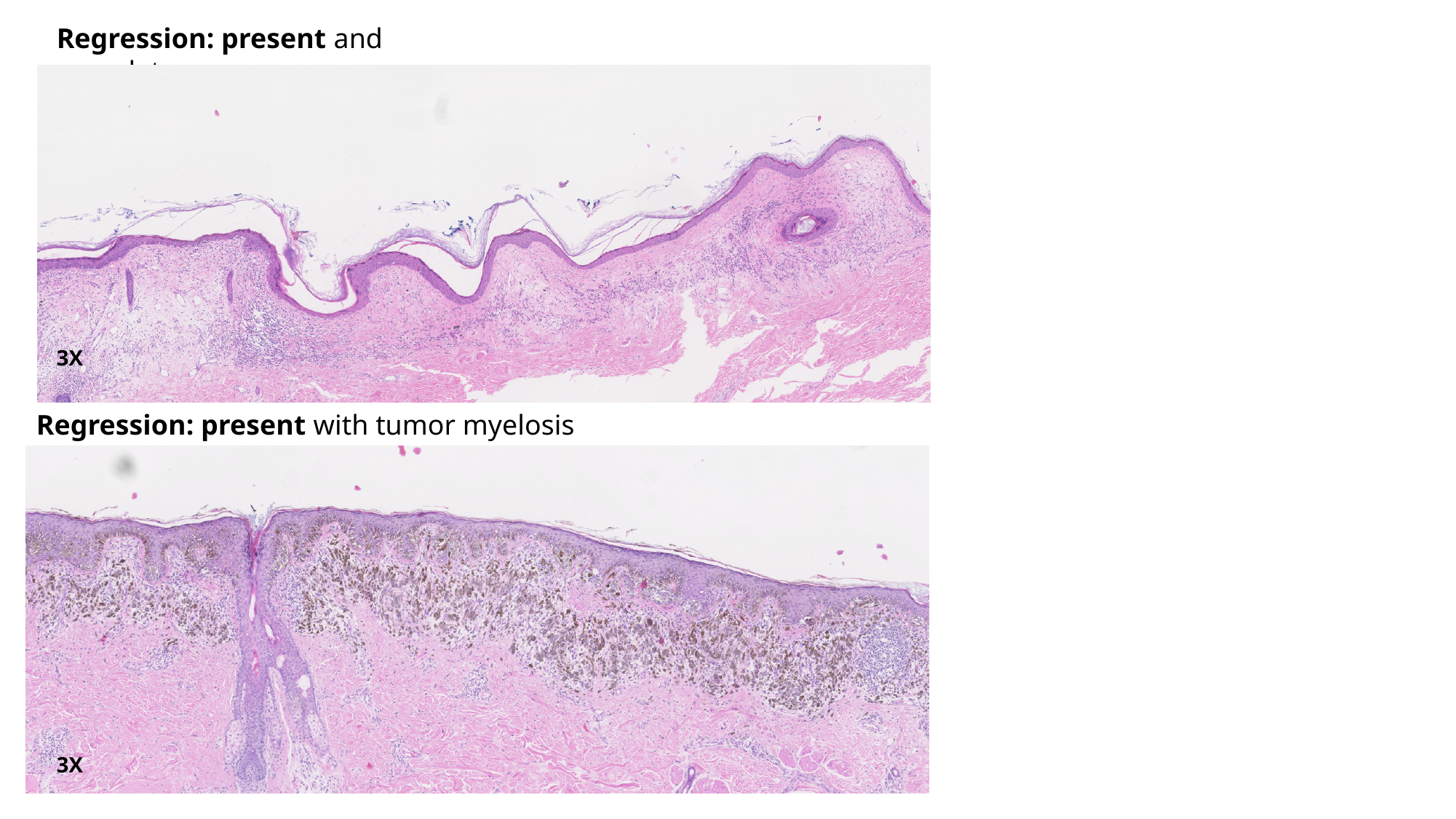

Regression: present and complete
3X
Regression: present with tumor myelosis
3X

### Slide 11
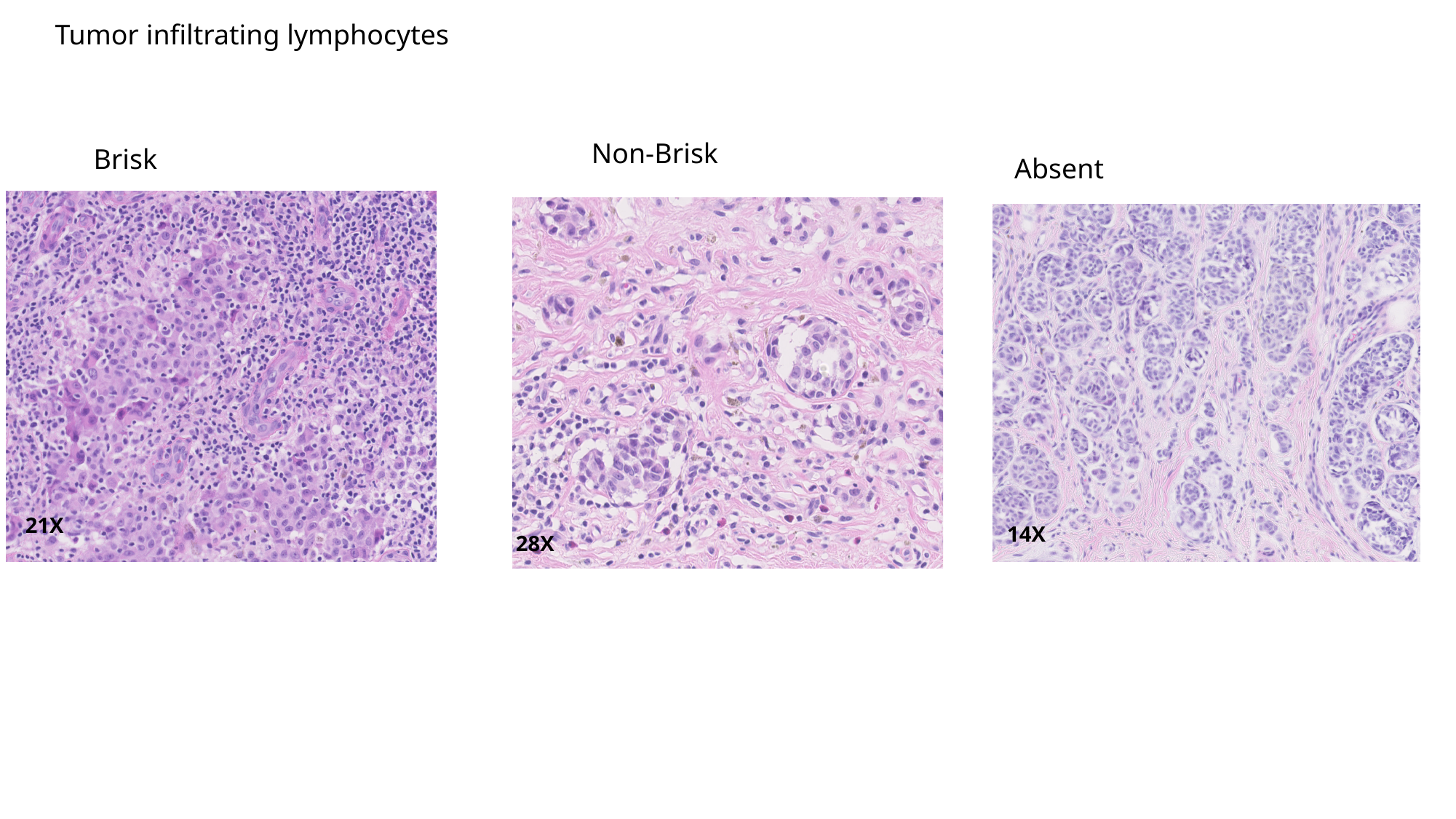

Tumor infiltrating lymphocytes
Non-Brisk
Brisk
Absent
21X
14X
28X

### Slide 12
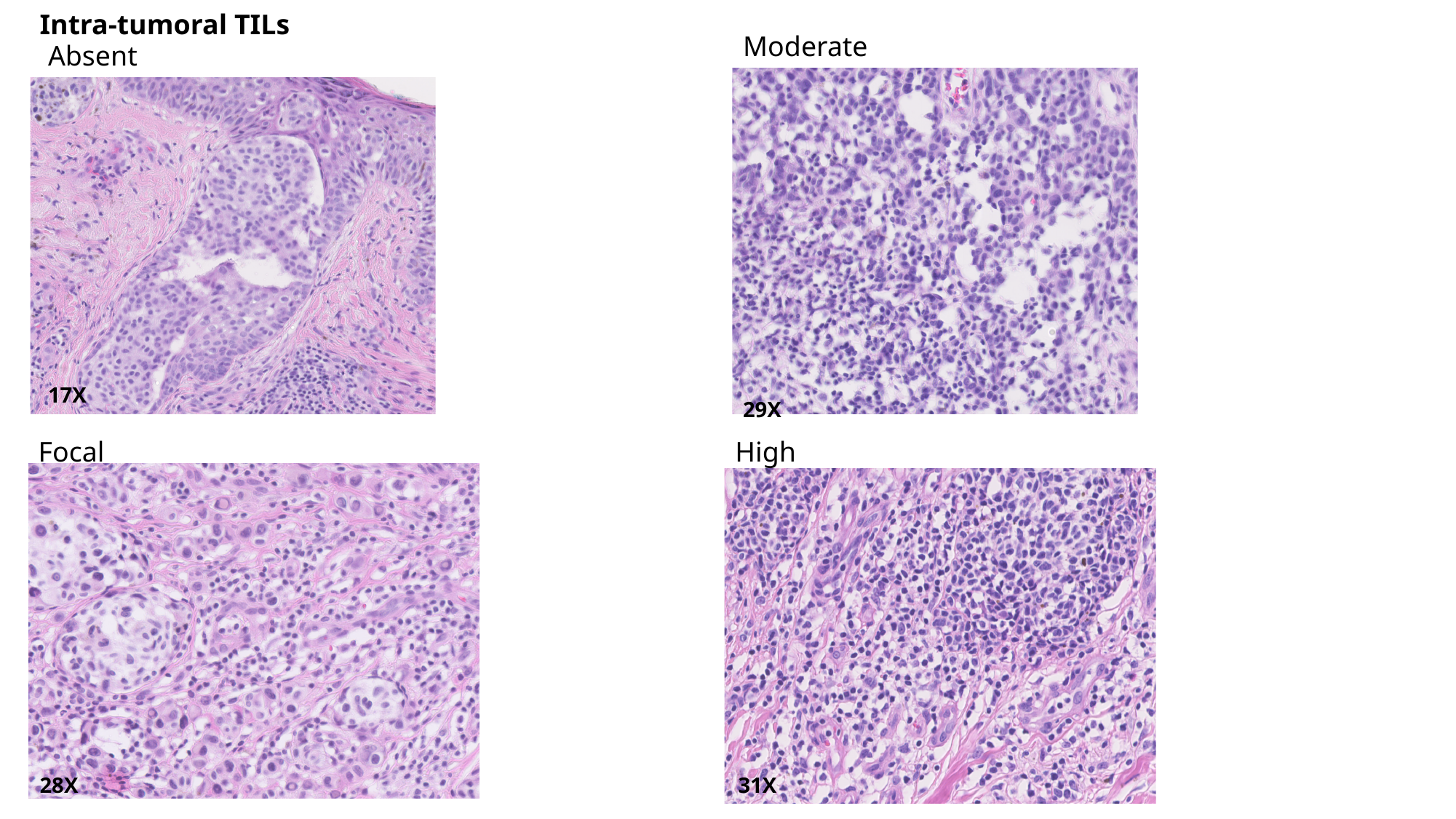

Intra-tumoral TILs
Moderate
Absent
17X
29X
Focal
High
31X
28X

### Slide 13
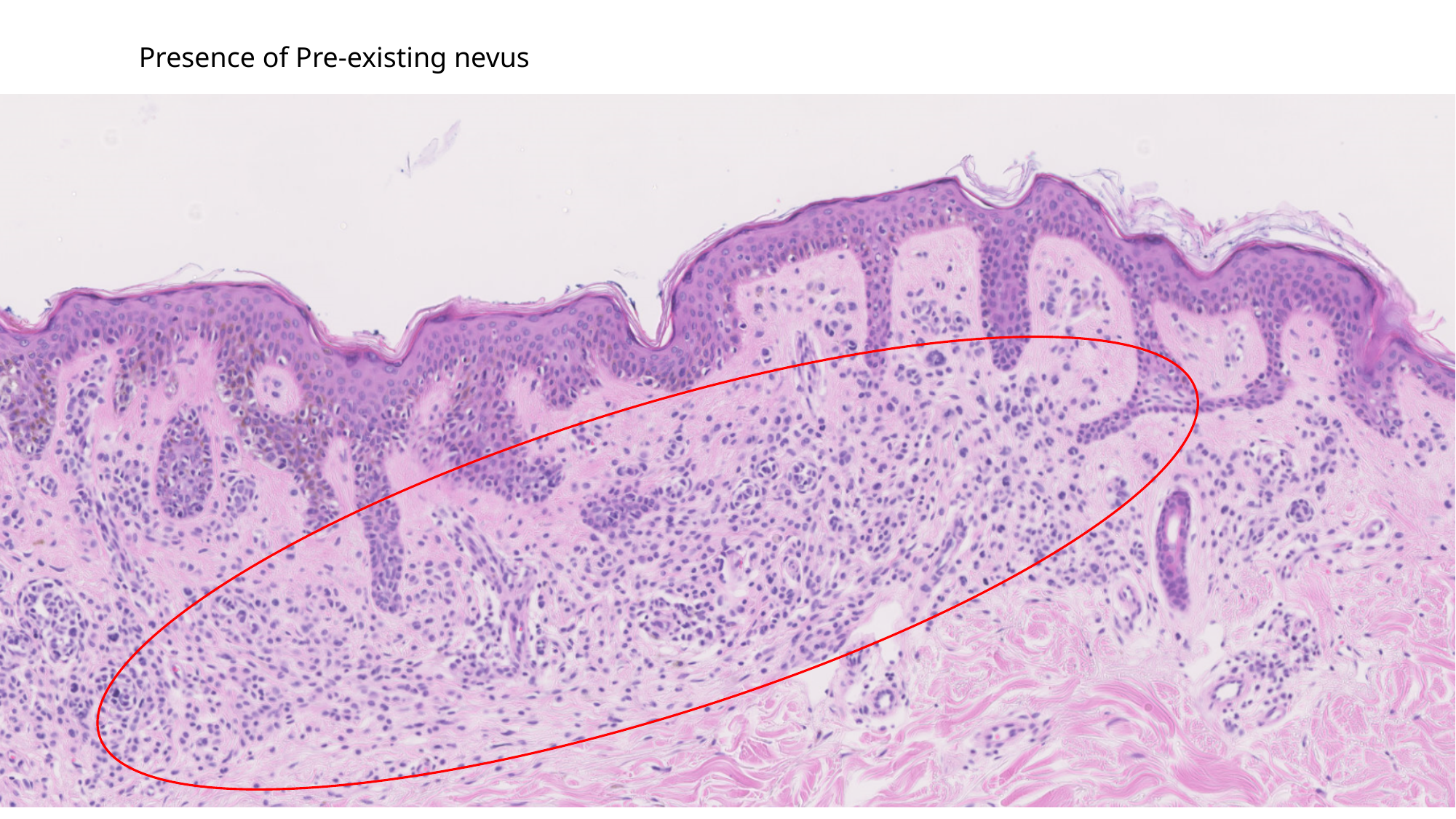

Presence of Pre-existing nevus

### Slide 14
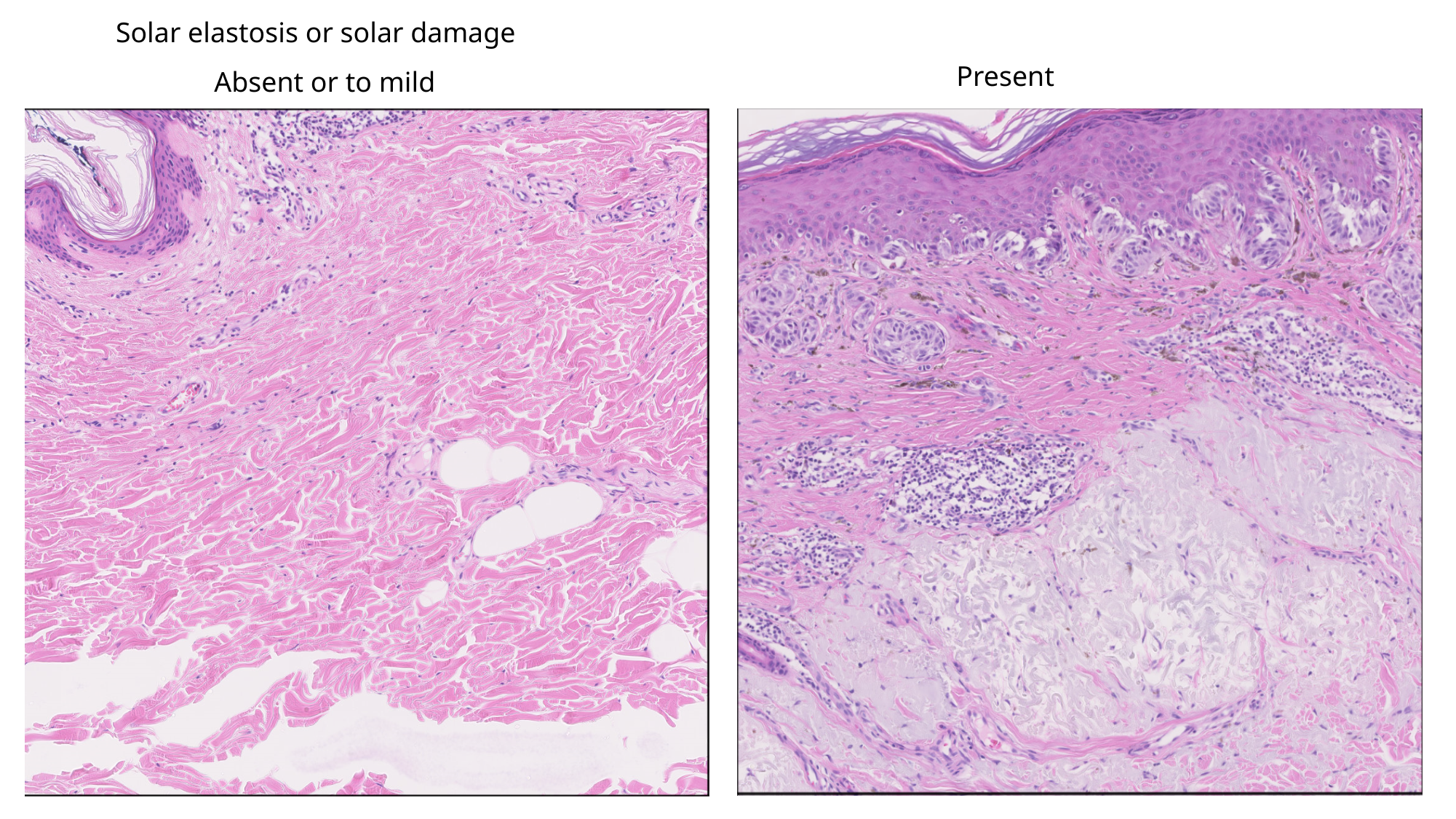

Solar elastosis or solar damage
Present
Absent or to mild

### Slide 15
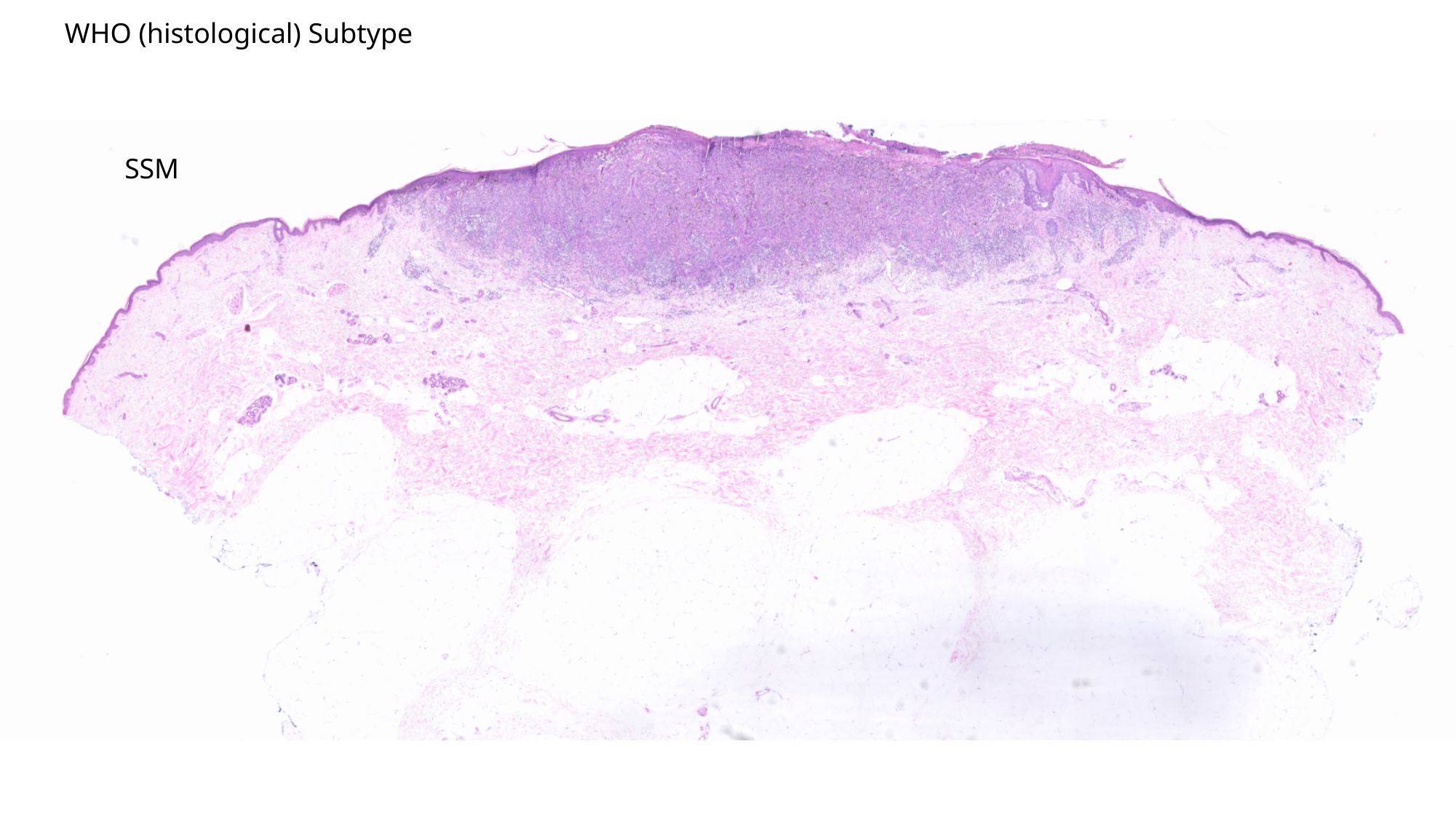

WHO (histological) Subtype
SSM

### Slide 16
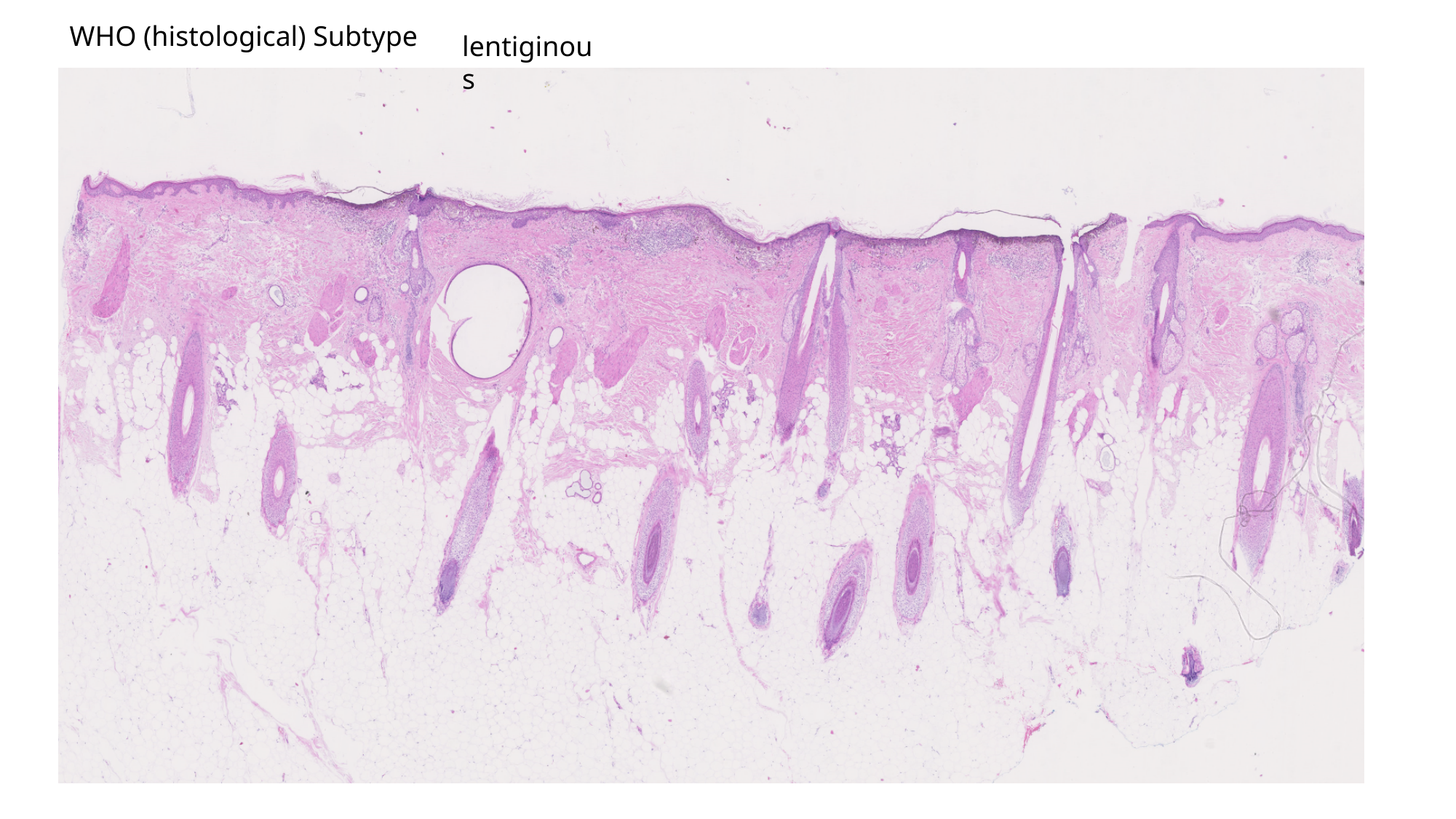

lentiginous
WHO (histological) Subtype

### Slide 17
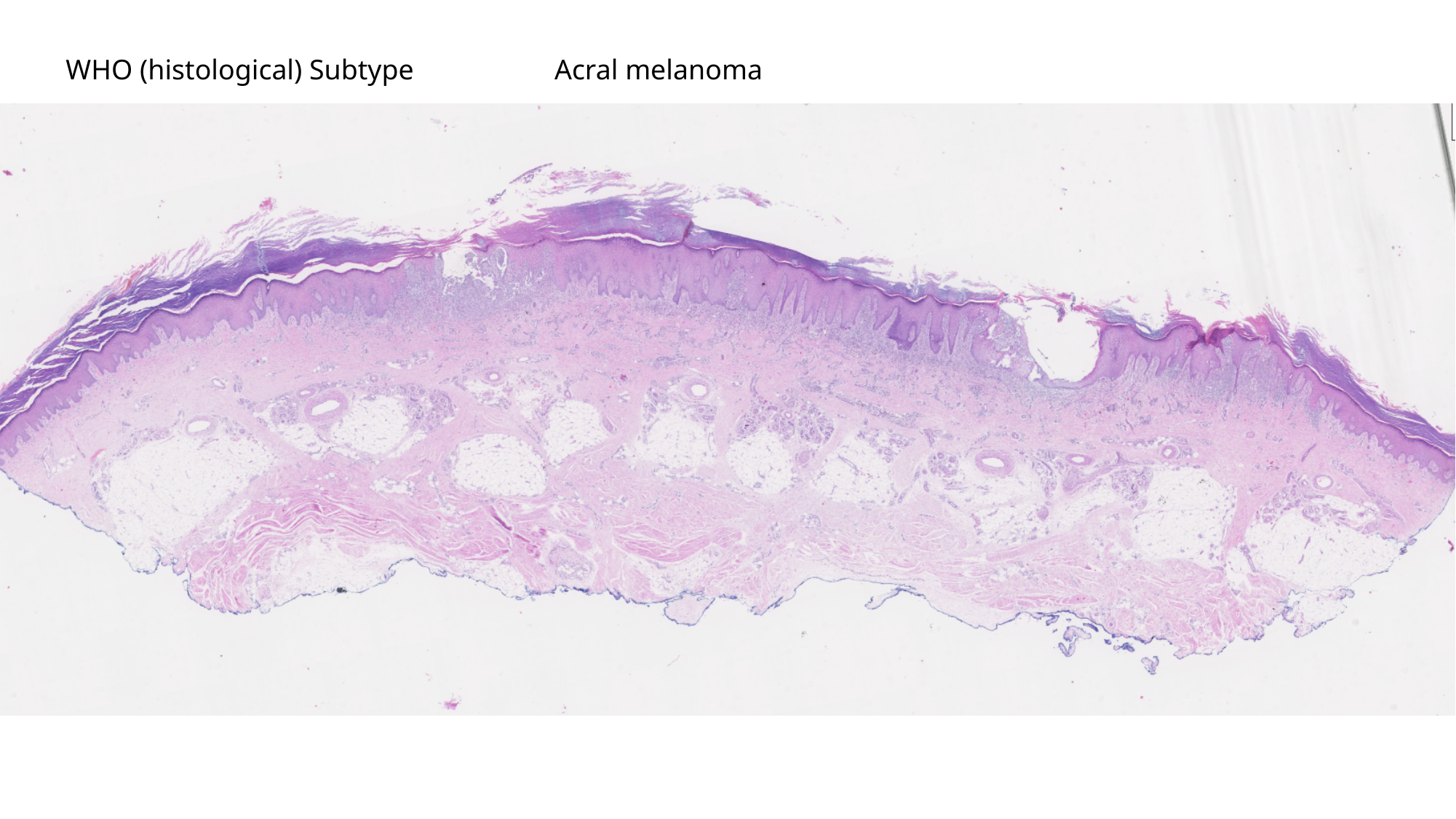

WHO (histological) Subtype
Acral melanoma

### Slide 18
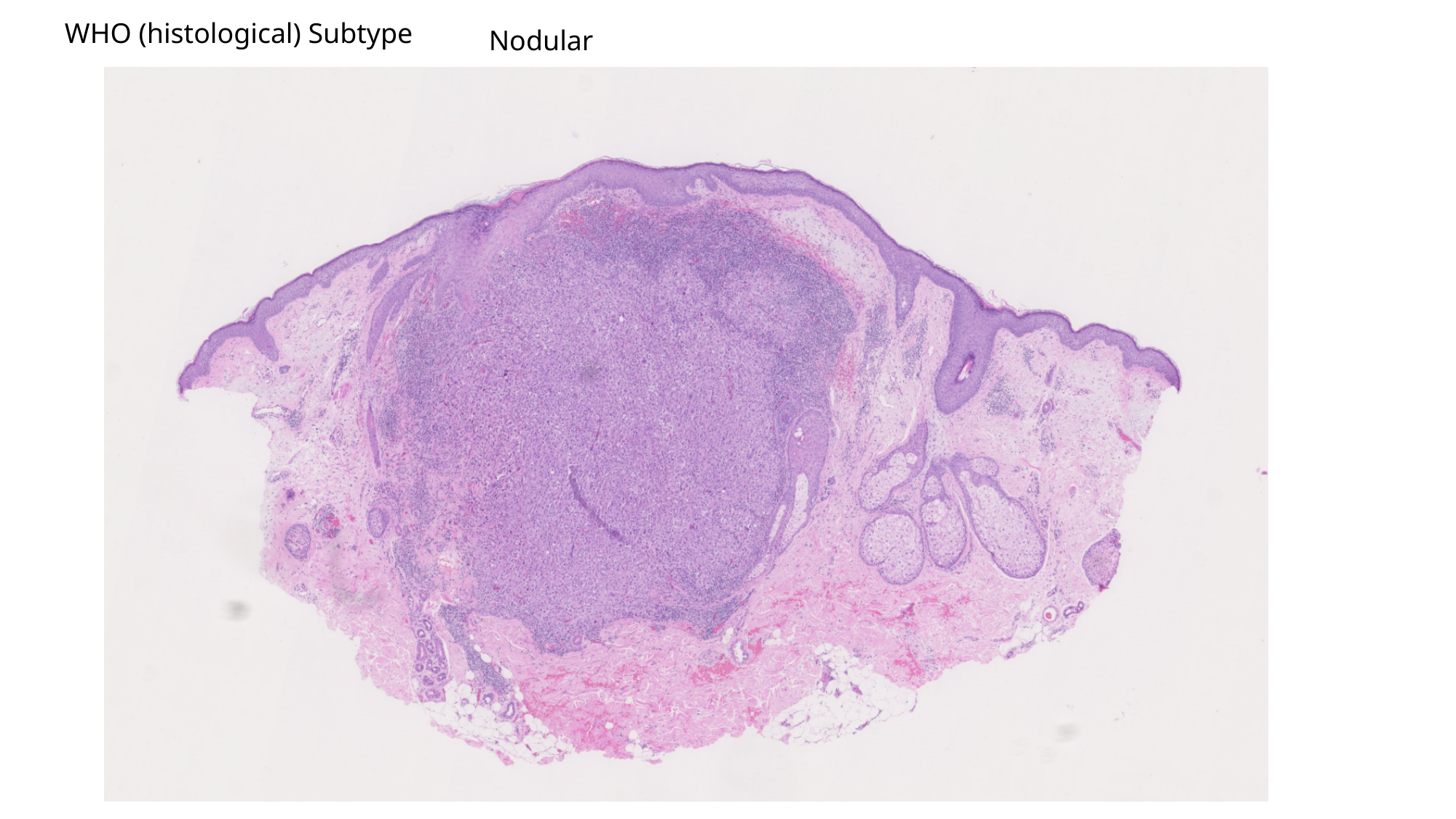

WHO (histological) Subtype
Nodular

### Slide 19
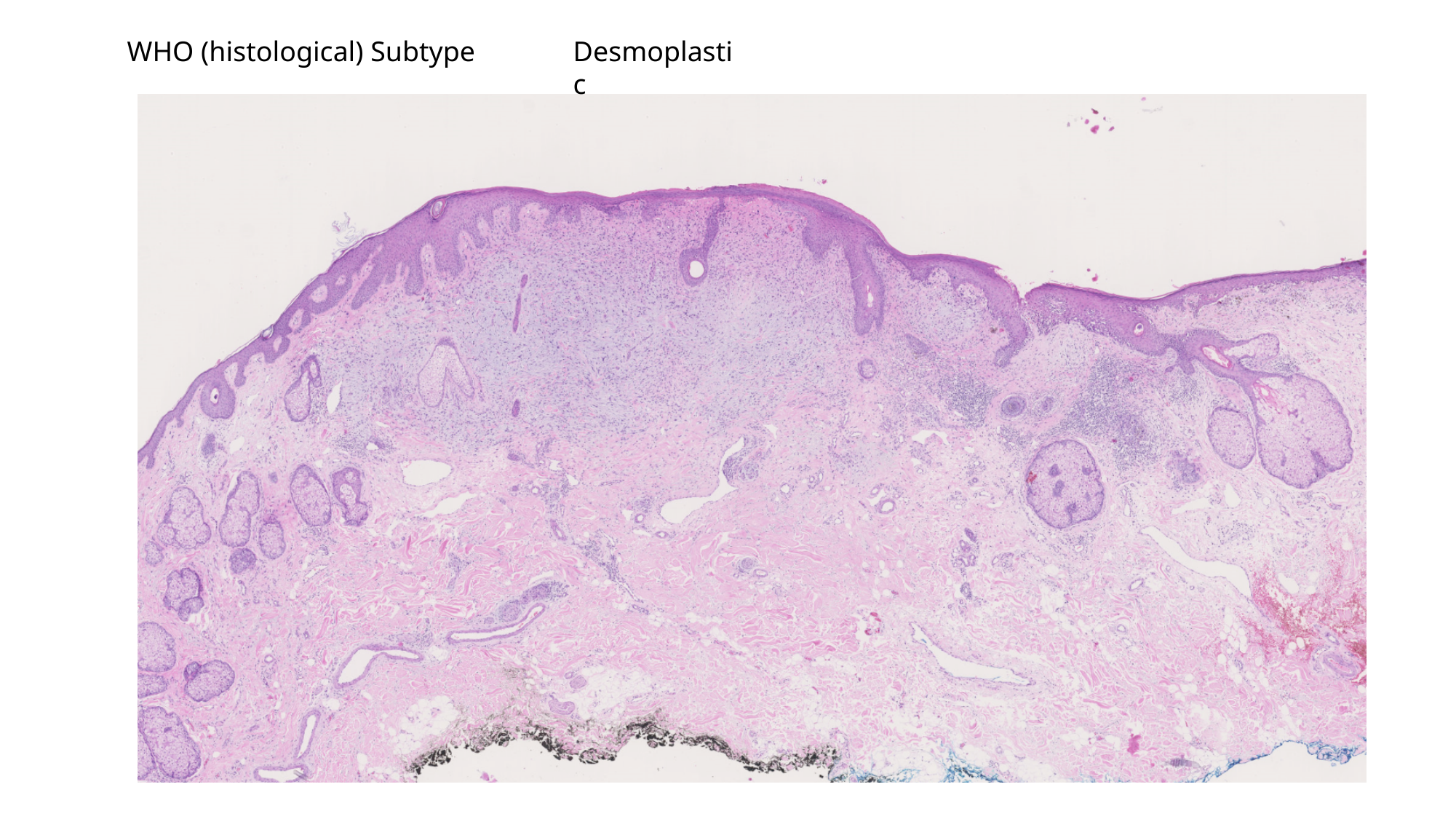

WHO (histological) Subtype
Desmoplastic

### Slide 20
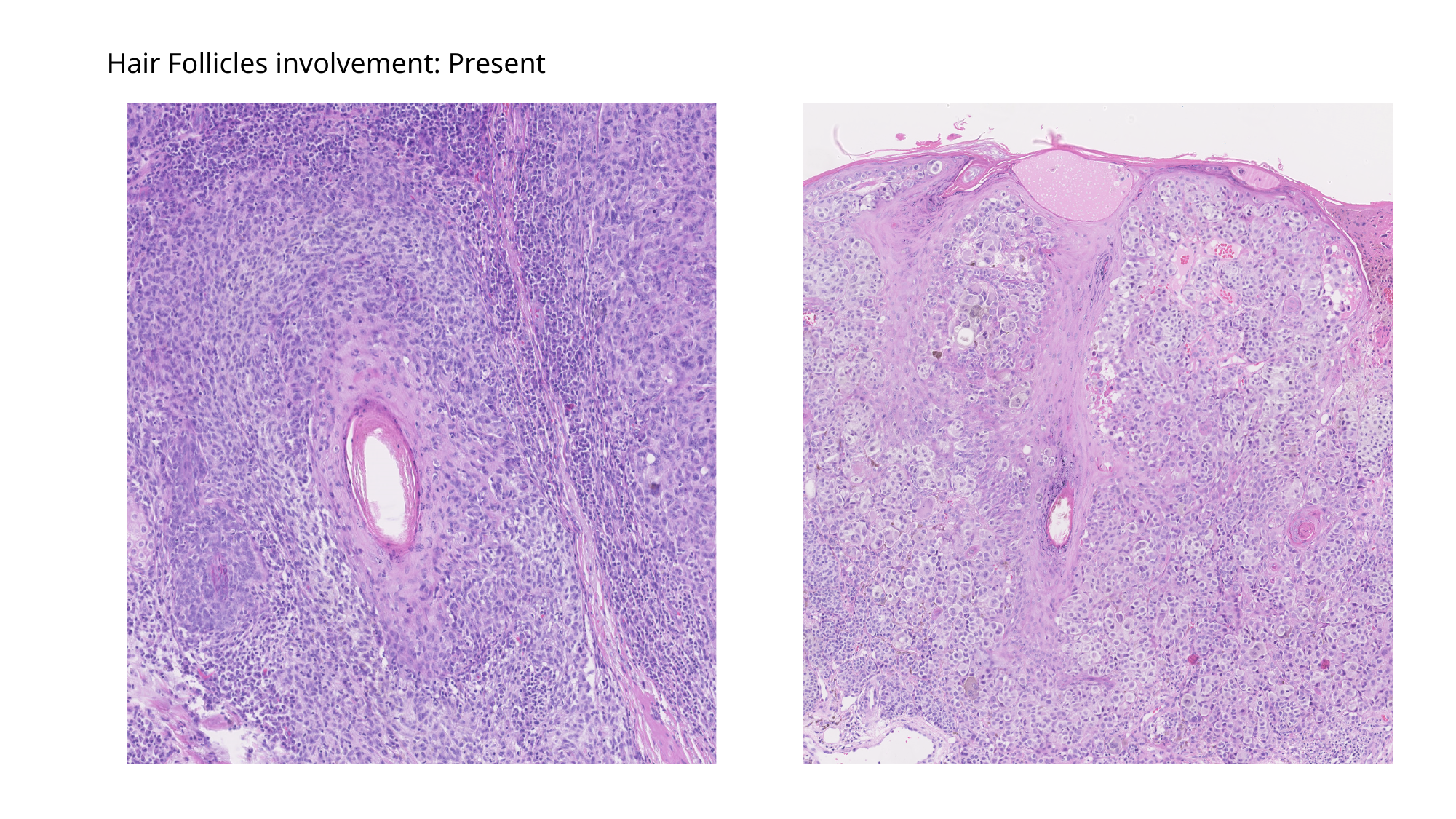

Hair Follicles involvement: Present
