## Supplementary material for "Automated histopathological measurements of the tumor micro-environment predict distant metastasis after stage I/II Melanoma: discovery and validation in the population-based Dutch Early-Stage Melanoma (D-ESMEL) study": all supplementary tables and figures: Supplementary methods.docx

1. **Supplementary method 1: Identification of relevant histopathological variables from the literature**:

Search terms to identify relevant features for histopathological scoring and the list of studies that were identified as relevant:

**Search:**

All histopathology studies on hematoxylin and eosin slides in cutaneous melanoma. Search terms per database are shown below the table.

| **Database searched** | **via** | **Years of coverage** | **Records** | **Records after duplicates removed** |
| --- | --- | --- | --- | --- |
| Embase | Embase.com | 1971 - Present | 560 | 376 |
| Medline ALL | Ovid | 1946 - Present | 420 | 420 |
| Web of Science Core Collection* | Web of Knowledge | 1975 - Present | 169 | 37 |
| Cochrane Central Register of Controlled Trials | Wiley | 1992 - Present | 9 | 0 |
| **Total** | | | **1158** | **833** |

*Science Citation Index Expanded (1975-present) ; Social Sciences Citation Index (1975-present) ; Arts & Humanities Citation Index (1975-present) ; Conference Proceedings Citation Index- Science (1990-present) ; Conference Proceedings Citation Index- Social Science & Humanities (1990-present) ; Emerging Sources Citation Index (2015-present)

**embase.com	570**

('cutaneous melanoma'/de OR 'primary melanoma'/de OR 'metastatic melanoma'/de OR (melanoma/de AND 'skin tumor'/exp) OR ((cutan* OR skin* OR derma* OR primar* OR metasta*) NEAR/6 melano*):Ab,ti) AND (histopathology/de OR histology/de OR pathology/de OR classification/de OR 'observer variation'/de OR (histopatholog* OR histomorpholog* OR histolog* OR patholog* OR classification* OR characteristic* OR feature* OR criteri* OR ((observer* OR interobserver* OR intraobserver*) NEAR/3 varia*)):Ab,ti) AND (hematoxylin/de OR eosin/de OR 'hematoxylin and eosin stain'/de OR 'hematoxylin and eosin staining'/de OR 'hematoxylin eosin stain'/de OR 'hematoxylin eosin staining'/de OR (hematoxylin* OR haematoxylin* OR eosin OR h-e):Ab,ti) NOT ('epiluminescence microscopy'/mj OR microscopy/mj OR 'mucosal melanoma'/mj OR 'uvea melanoma'/exp/mj OR 'spitz nevus'/de OR nevus/mj OR (dermoscop* OR microscop* OR ((mucos* OR uvea*) NEAR/3 melanom*) OR spitz* OR nevus* OR nevi OR naevus* OR naevi OR (lymphat* NEAR/6 sentinel*) OR non-melanom*):ti) NOT ([animals]/lim NOT [humans]/lim)

**Medline ALL**

(Skin Neoplasms/ OR (Melanoma/ AND exp Skin Neoplasms/) OR ((cutan* OR skin* OR derma* OR primar* OR metasta*) ADJ6 melano*).ab,ti.) AND (Histology/ OR Pathology/ OR Classification/ OR Observer Variation/ OR (histopatholog* OR histomorpholog* OR histolog* OR patholog* OR classification* OR characteristic* OR feature* OR criteri* OR ((observer* OR interobserver* OR intraobserver*) ADJ3 varia*)).ab,ti.) AND (Hematoxylin/ OR Eosine Yellowish-YS/ OR (hematoxylin* OR haematoxylin* OR eosin OR h-e).ab,ti.) NOT (* Dermoscopy/ OR * Microscopy/ OR "Nevus, Epithelioid and Spindle Cell"/ OR * Nevus/ OR (dermoscop* OR microscop* OR ((mucos* OR uvea*) ADJ3 melanom*) OR spitz* OR nevus* OR nevi OR naevus* OR naevi OR (lymphat* ADJ6 sentinel*) OR non-melanom*).ti.) NOT (exp animals/ NOT humans/)

**Web of Science Core Collection***

TS=((((cutan* OR skin* OR derma* OR primar* OR metasta*) NEAR/5 melano*)) AND ((histopatholog* OR histomorpholog* OR histolog* OR patholog* OR classification* OR characteristic* OR feature* OR criteri* OR ((observer* OR interobserver* OR intraobserver*) NEAR/2 varia*))) AND ((hematoxylin* OR haematoxylin* OR eosin OR h-e)) NOT ((dermoscop* OR microscop* OR ((mucos* OR uvea*) NEAR/2 melanom*) OR spitz* OR nevus* OR nevi OR naevus* OR naevi OR (lymphat* NEAR/5 sentinel*) OR non-melanom*)))

**Cochrane Central Register of Controlled Trials**

(((cutan* OR skin* OR derma* OR primar* OR metasta*) NEAR/6 melano*):Ab,ti) AND ((histopatholog* OR histomorpholog* OR histolog* OR patholog* OR classification* OR characteristic* OR feature* OR criteri* OR ((observer* OR interobserver* OR intraobserver*) NEAR/3 varia*)):Ab,ti) AND ((hematoxylin* OR haematoxylin* OR eosin OR h NEXT e):Ab,ti) NOT ((dermoscop* OR microscop* OR ((mucos* OR uvea*) NEAR/3 melanom*) OR spitz* OR nevus* OR nevi OR naevus* OR naevi OR (lymphat* NEAR/6 sentinel*) OR non NEXT melanom*):ti) NOT (animals NOT humans)

Histopathological measurements description table:

| **Histological Feature** | **Reference** |
| --- | --- |
| Epidermal contour | 1 |
| Pagetoid spread | 1 |
| Nest formation of intraepidermal melanocytes | 1 |
| Mitotic index | 2,5 |
| Regression | 2 |
| Tumor infiltrating lymphocytes (TILs score) | 2,3,5 |
| stromal TILs% | 3 |
| intratumoral TILs | 3 |
| pre-existing nevus | 4, 6 |
| Solar damage | 4, 6 |
| WHO subtype | 1,5,6 |
| Clark level | 5 |
| Satellitosis | 5 |
| Perineural invasion | 5 |
| hair follicle involvement | 5 |
| plasma cell cluster | 3 |

List of studies (references):

1. Viros A, Fridlyand J, Bauer J, Lasithiotakis K, Garbe C, Pinkel D, Bastian BC. Improving melanoma classification by integrating genetic and morphologic features. PLoS Med. 2008 Jun 3;5(6):e120. doi: 10.1371/journal.pmed.0050120. PMID: 18532874; PMCID: PMC2408611.
2. Beer J, Xu L, Tschandl P, Kittler H. Growth rate of melanoma in vivo and correlation with dermatoscopic and dermatopathologic findings. *Dermatol Pract Concept*. 2011;1(1):59-67. Published 2011 Jan 31. doi:10.5826/dpc.dp0101a13
3. Mihm, M. C. & Mulé, J. J. Reflections on the Histopathology of Tumor-Infiltrating Lymphocytes in Melanoma and the Host Immune Response. *Cancer Immunol. Res.* **3**, 827–835 (2015).
4. Landi MT, Bauer J, Pfeiffer RM, Elder DE, Hulley B, Minghetti P, Calista D, Kanetsky PA, Pinkel D, Bastian BC. MC1R germline variants confer risk for BRAF-mutant melanoma. Science. 2006 Jul 28;313(5786):521-2. doi: 10.1126/science.1127515. Epub 2006 Jun 29. PMID: 16809487.
5. Tejera-Vaquerizo A, Fernández-Figueras MT, Santos-Briz A, Ríos-Martín JJ, Monteagudo C, Fernández-Flores A, Requena C, Traves V, Descalzo-Gallego MA, Rodríguez-Peralto JL. Protocol for the Histologic Diagnosis of Cutaneous Melanoma: Consensus Statement of the Spanish Society of Pathology and the Spanish Academy of Dermatology and Venereology (AEDV) for the National Cutaneous Melanoma Registry. Actas Dermosifiliogr (Engl Ed). 2021 Jan;112(1):32-43. English, Spanish. doi: 10.1016/j.ad.2020.09.002. Epub 2020 Oct 7. PMID: 33038295; PMCID: PMC7540207.
6. Scolyer RA, Long GV, Thompson JF. Evolving concepts in melanoma classification and their relevance to multidisciplinary melanoma patient care. Mol Oncol. 2011;5(2):124-136. doi:10.1016/j.molonc.2011.03.002
2. **Supplementary method 2: Testing the agreement between the two pathologists in the validation cohort for 80 samples :** Interobserver agreement for Stromal TILS was accessed using the interclass correlation coefficient (ICC), as this was a continues variable. Agreement for categorical variables (pagetoid spread and hair follicle involvement) was evaluated using the Cohen’s Kappa Statistic (Cohen’s κ). The agreement between the pathologists ensured the reproducibility of the scoring system used in this study. ICC >0.75 was considered excellent agreement, ICC of 0.4 - 0.75 was considered as moderate agreement, while ICC<0.4 was considered as poor agreement. For the Cohen’s Kappa , a value of <0.20 indicate poor agreement, 0.21–0.40 indicated a fair agreement, 0.41- 0.60 indicate moderate agreement, 0.61–0.80 indicate good agreement, and values >0.80 indicate very good agreement. The results are shown below:

| Variable | Statistic | Value (95% CI) | N | Interpretation |
| --- | --- | --- | --- | --- |
| Stromal TILs | ICC | 0.82 (0.69–0.89) | 76 | Very Good |
| Pagetoid spread | Cohen’s κ | 0.67 (0.49–0.85) | 79 | Substantial |
| Hair follicle involvement | Cohen’s κ | 0.70 (0.68–0.85) | 80 | Substantial |

3. **Supplementary method 3 : Multiple imputation with using chained equation and backward selection to identify independent predictors with manual histopathological variables:**

Missing data in the pathological scoring occurred when pathologist was unable to score the features due to the low quality H&E images or missing tissue section if it was a biopsy. Therefore, we assume that values were missing at random and were imputed with multiple imputation using chained equation (*mice* R package (version 3.19.0). The imputation model included all the variables presented in the Supplementary table 2**,** as well as the metastatic outcome. Ten imputed datasets were created using 30 iterations. Coverage plots were used to visualize the convergence of the mice algorithm. For the automated feature data, the segmentation output for each image was reviewed by two expert pathologists as previously described [25], and samples with missing data due to poor segmentation quality were excluded from the analysis.

To identify the best combination of histological features that predict metastasis risk using the manual pathological scoring or the automated features extraction, we performed multivariable conditional logistic regression analyses. The dependent variable was the occurrence of distant metastasis. The initial models for each data included all the measured features, and backward variable selection was applied to keep only the predictive features. The pathological scoring data was imputed and therefore multiple datasets from the multiple imputation needed to be combined to perform the variable selection. For this purpose, we used a stepwise selection method [26]. First, for each imputed dataset, we performed a stepwise variable selection procedure using the Akaike’s Information as the selection criterion. then, we recorded how frequently each variable was selected across the imputed datasets. Finaly, variables that were frequently selected were evaluated using Wald test to determine whether they should be retained in the final model.

4. **Supplementary method 4: Performance of the existing risk models in melanoma:**

The cases and controls in the discovery set of the D-ESMEL study are matched for well-known prognostic variables: Breslow thickness, ulceration, age and sex. However, other validated prognostic models also include additional variables. To test the prognostic value of those variables in addition to our matching variables, we constructed multivariable models incorporating variables used in two publicly available calculators developed by the Melanoma Institute Australia (MAI). First, we tested features from Thin Melanoma recurrence risk model [28]. For patients with T1 melanoma from the discovery set, we evaluated, anatomic location, histological subtype, and sentinel lymph node (SLN) status. Age, sex, Breslow thickness and ulceration are also included in the MIA model, but not in our model, because we already matched on those factors. Secondly, we tested the features included in the Stage II melanoma survival risk model [29]. Briefly, for Stage II patients from the discovery set, we included, histological subtype, location, mitotic rate, satellite lesions, lymphovascular invasion, tumor-infiltrating lymphocytes (TILs), regression, and SLN status. Age, sex, ulceration, Breslow thickness were matched and not included in the model.
