## Supplementary figures and images for "Automated histopathological measurements of the tumor micro-environment predict distant metastasis after stage I/II Melanoma: discovery and validation in the population-based Dutch Early-Stage Melanoma (D-ESMEL) study"

### Supplimentary Figure 1.pdf

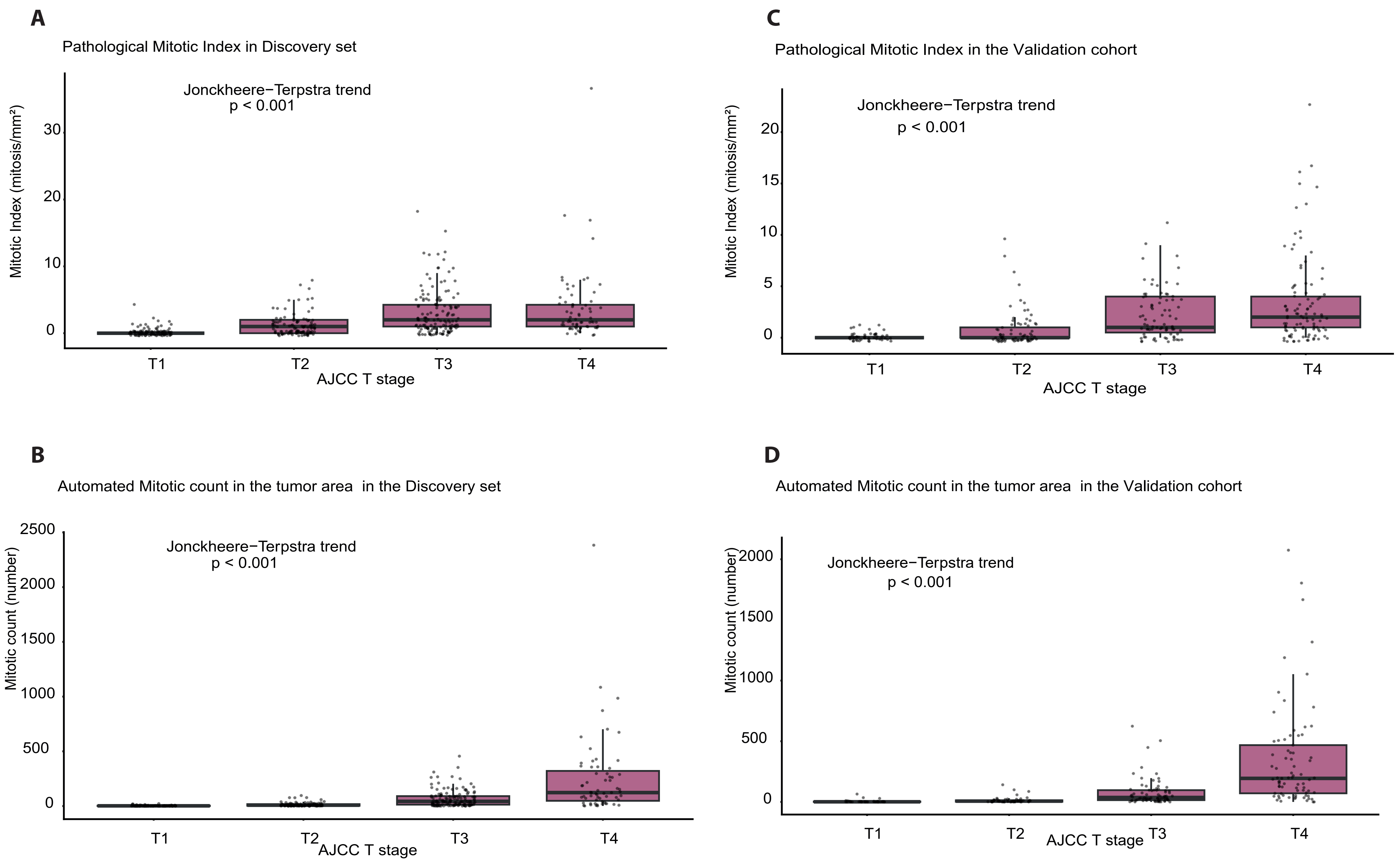

### Supplimentary Figure 2.pdf

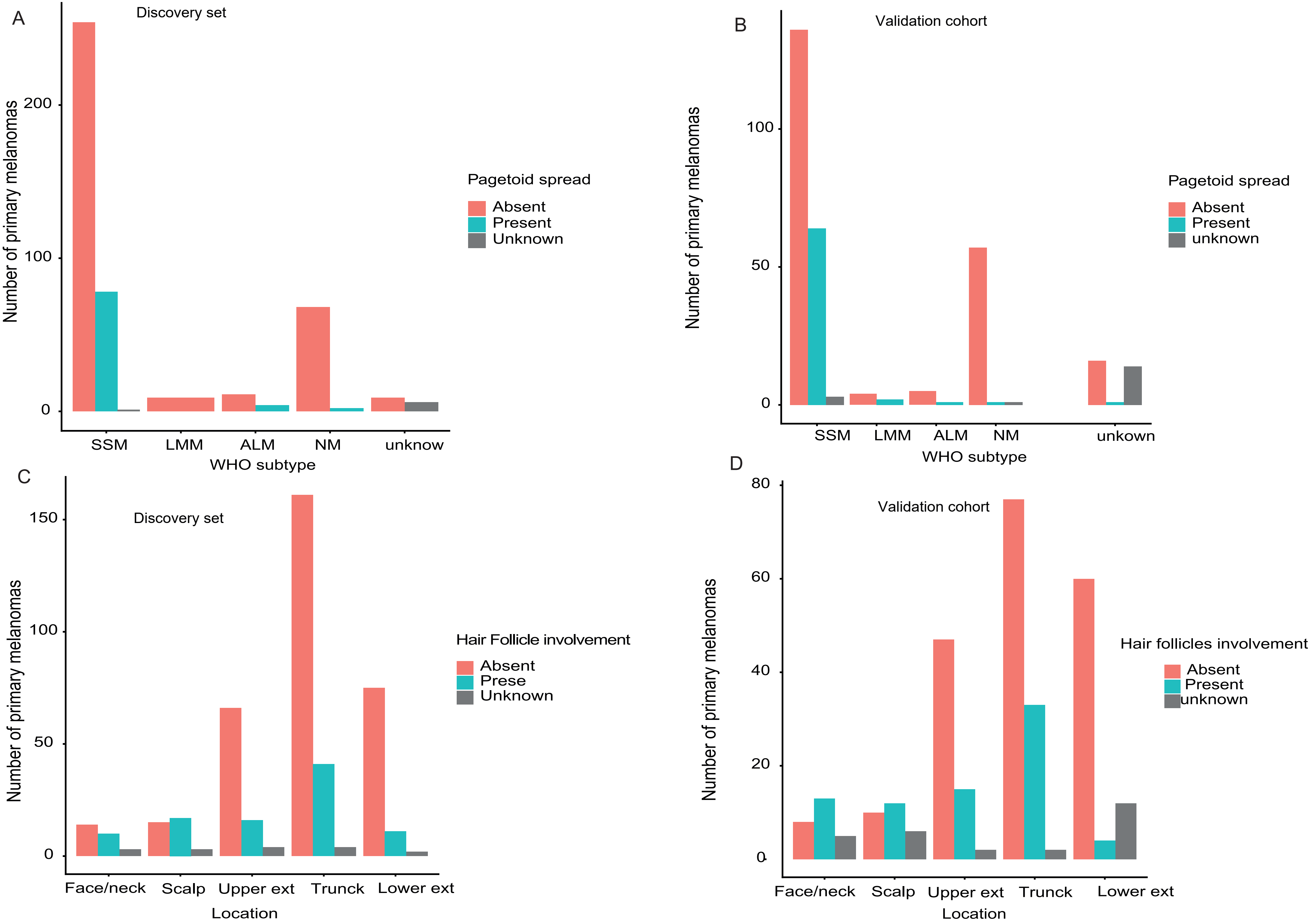

### Supplimentary Figure 3.pdf

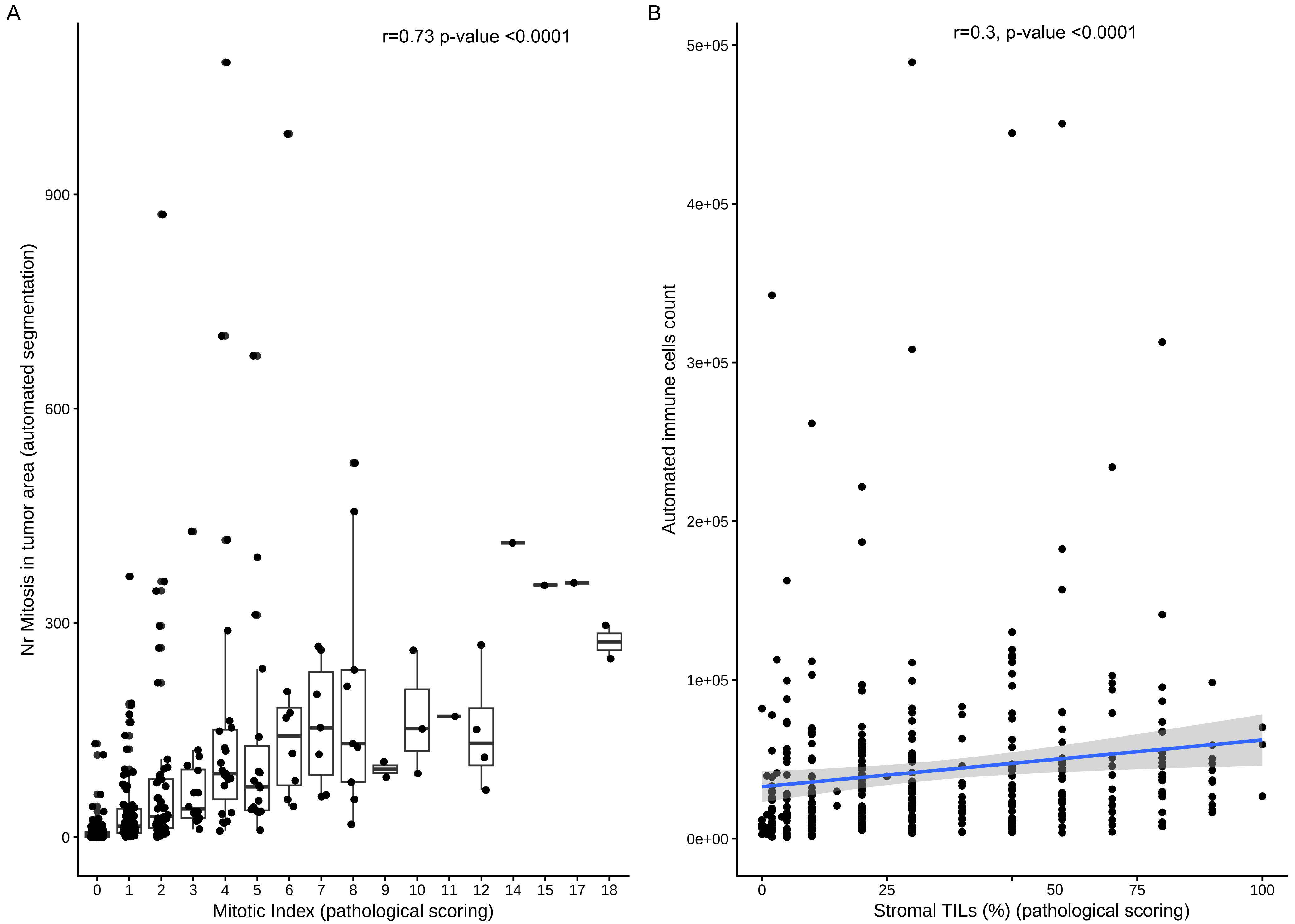
